# Large language models for extracting histopathologic diagnoses of colorectal cancer and dysplasia from electronic health records

**DOI:** 10.1101/2024.11.27.24318083

**Authors:** Brian Johnson, Tyler Bath, Xinyi Huang, Mark Lamm, Ashley Earles, Hyrum Eddington, Anna M. Dornisch, Lily J. Jih, Samir Gupta, Shailja C. Shah, Kit Curtius

**Affiliations:** Division of Biomedical Informatics, Department of Medicine, University of California San Diego, La Jolla, CA, USA; Veterans Medical Research Foundation, San Diego, CA, USA; VA San Diego Healthcare System, San Diego, CA, USA; Department of Pathology, University of California San Diego, La Jolla, CA, USA; Division of Gastroenterology, University of California San Diego, La Jolla, CA, USA; Department of Radiation Medicine & Applied Sciences, University of California San Diego, La Jolla, CA; Moores Cancer Center, University of California San Diego, La Jolla, CA, USA

**Keywords:** artificial intelligence, large language models, natural language processing, biomedical informatics, colorectal cancer, inflammatory bowel disease

## Abstract

**Background:** Accurate data resources are essential for impactful medical research, but available structured datasets are often incomplete or inaccurate. Recent advances in open-weight large language models (LLMs) enable more accurate data extraction from unstructured text in electronic health records (EHRs) but have not yet been thoroughly validated for challenging diagnoses such as inflammatory bowel disease (IBD)-related neoplasia.

**Objective:** Create a validated approach using LLMs for identifying histopathologic diagnoses in pathology reports from the nationwide Veterans Health Administration database, including patients with genotype data within the Million Veteran Program (MVP) biobank.

**Design:** Our approach utilizes simple ‘yes/no’ question prompts for following phenotypes of interest: any colorectal dysplasia, high-grade dysplasia and/or colorectal adenocarcinoma (HGD/CRC), and invasive CRC. We validated the method on diagnostic tasks by applying prompts to reports from patients with IBD (and validated separately in non-IBD) and calculated F-1 scores as a balanced accuracy measure.

**Results:** In patients with IBD in MVP, we achieved F1-scores of 96.1% (95% CI 92.5-99.4%) for identifying dysplasia, 93.7% (88.2-98.4%) for identifying HGD/CRC, and 98% (96.3-99.4%) for identifying CRC. In patients without IBD in MVP, we achieved F1-scores of 99.2% (98.2-100%) for identifying any colorectal dysplasia, 96.5% (93.0-99.2%) for identifying HGD/CRC, and 95% (92.8-97.2%) for identifying CRC using LLM Gemma-2.

**Conclusion:** LLMs provided excellent accuracy in extracting the diagnoses of interest from EHRs. Our validated methods generalized to unstructured pathology notes, even withstanding challenges of resource-limited computing environments. This may therefore be a promising approach for other clinical phenotypes given the minimal human-led development required.

**Key Messages:** *What is already known on this topic:* Extracting structured data from free-text health records, such as pathology reports, remains a significant challenge in clinical research. Traditional natural language processing methods require extensive development and are often difficult to generalize across settings, limiting their usefulness for large-scale, reproducible data extraction.

*What this study adds:* This study demonstrates that relatively small (8-9 billion parameter) publicly available large language models can accurately extract cancer and dysplasia diagnoses from pathology reports without additional task-specific training or fine-tuning.

*How this study might affect research, practice or policy:* By enabling accurate data extraction from clinical text, large language models offer a scalable and accessible solution for structuring clinical data, reducing the burden of algorithm development and/or manual data curation. These advancements facilitate expanded access to high-quality real-world medical data for clinical and translational research.

## Introduction

The expected breakthroughs in personalized treatments and improved medical outcomes have yet to fully materialize despite the exponential increase in volume of healthcare data available for research. One obstacle impeding these advances is the quality and accessibility of the vast data generated and stored as part of usual healthcare.

As an example use case, tailoring colonoscopy screening ages and surveillance intervals based on accurate risk stratification informed by large, high-quality datasets has real potential to reduce both the incidence of colorectal cancer, as well as the number of unnecessary colonoscopies that add burden to both patients and the healthcare system^1^. Current risk stratification approaches in both the general population and those with inflammatory bowel disease (IBD) are based on few clinical variables, which are often associated with widely varied published estimates of risk^2,3^. For example, when lesions such as adenomas or flat low-grade dysplasia are diagnosed, current guidelines for all-comers recommend surveillance colonoscopy every 1-10 years (or 1-5 years in patients with IBD) based on clinical risk stratification^3,4^. As such, screening guidelines essentially serve as heuristics, representing the best approach with limited data^5^. Our overall goal is to improve the quality of available large-scale data resources—an essential prerequisite for accurate downstream analyses to improve personalized medicine—by leveraging artificial intelligence to extract clinical information, with a focus on histopathologic diagnoses in the present study.

Traditionally, “rules-based” natural language processing (NLP) algorithms have dominated structured data extraction in this area. Briefly, NLP translates natural language into data formats that are easier for computers to process and analyze^6^. Performance of these algorithms is excellent, with F1-scores frequently above 99% for identifying adenomas in data from the general population^7–14^. Alternatively, published deep-learning or embedding-based models are less common for classifying pathology findings, though work by Syed et al. reported an F1-score of 95% for identifying neoplastic (dysplastic) polyps^15^. However, there are many drawbacks to current approaches. Development requires extensive human effort to refine algorithms: creating concepts (e.g., enumerating all possible ways that each diagnosis could be written), identifying negation (ensuring that expressions of the absence or uncertainty of a diagnosis are captured and related to the correct concept), associating terms with their respective anatomical locations, and modifying the algorithm to address “edge” cases. Adapting these algorithms to new use cases or different databases presents similar challenges, as development is often tailored to the formatting and style of a specific hospital system, patient cohort, and/or time period^16^.

Previous NLP approaches for such diagnostic tasks have not been tested in IBD populations specifically and may not be appropriate for use in the setting of IBD. For example, pathology reports from patients with IBD are often slightly different from reports from the non-IBD population. In the IBD patient cohort, historical terminology may be present (e.g., Dysplasia Associated Lesion/Mass or “DALMs”) and there are more instances of negation where pathologists explicitly rule out dysplasia and carcinoma than in patients without IBD. Additionally, few of these NLP approaches have been rigorously tested in identifying varying severities of dysplasia (e.g., low-grade, high-grade) and adenocarcinoma in either IBD or non-IBD. Thus, previous methods are largely insufficient for these examples and manual chart review of millions of patient records is infeasible in terms of human labor required. Consequently, inaccurate data or lack of large-scale data availability can have negative effects on patient outcomes, for example through misinformed screening guidelines that may miss deadly cancers.

As opposed to traditional NLP methods, large language models (LLMs) are capable of many tasks “out of the box” without additional tedious human-led development. Because of this, LLMs should be less susceptible to differences in formatting and style, working better across settings^17^. For example, applying LLMs to determine colonoscopy follow-up time recommendations that are in line with established guidelines has been shown to be feasible without task-specific training^18^. Recently, there have also been remarkable advances in the quality of open-weight LLMs released under permissive licenses. Open-weight models avoid critical legal and regulatory issues, allowing researchers to conduct inference without significant privacy risks. Specifically, these models can be uploaded to the same computing environment where the data is stored, enabling researchers to structure data without it leaving this secured space. Because these models do not undergo training or fine-tuning within the computing environment, there is no risk of them ‘remembering’ patient information or accidental data breaches. Moreover, using these models does not involve sending data to third parties, avoiding the associated logistic and privacy challenges. While current LLMs require significant computational bandwidth, they are rapidly becoming viable alternatives for large-scale applications as their efficiency improves.

Here, our study objectives were to test and compare the performance of LLMs, without any task-specific training, on their ability to extract and characterize the presence vs. absence of dysplasia and adenocarcinoma from unstructured colonoscopy-associated pathology reports. We hypothesized that an LLM approach would be straightforward to implement and achieve greater accuracy than previously published historic methods on similar tasks both in IBD and non-IBD populations. We show that LLMs, even in resource-limited environments, are accurate at identifying features from pathology reports in a way that is easily reproducible.

## Materials and Methods

As a brief overview, the framework described below first uses simple search terms to filter colonoscopy pathology reports to identify those that were potentially diagnostic for each of the three following concepts in the colon or rectum: any dysplasia, high-grade dysplasia or adenocarcinoma (HGD/CRC), and invasive adenocarcinoma (CRC). For IBD, we also considered a task for indefinite for dysplasia (IND). For each task, we posed a yes-or-no question to a large language model (LLM) to determine whether the report contained a diagnosis of that concept (Figure 1). We iteratively developed specific prompts for each concept as necessary and then validated the model performance in a separate corpus of pathology reports. The study population was divided into IBD and non-IBD, and model validation was performed independently in both populations. We used stratified sampling for validation to account for the potentially low prevalence of some concepts.

**Figure 1.**
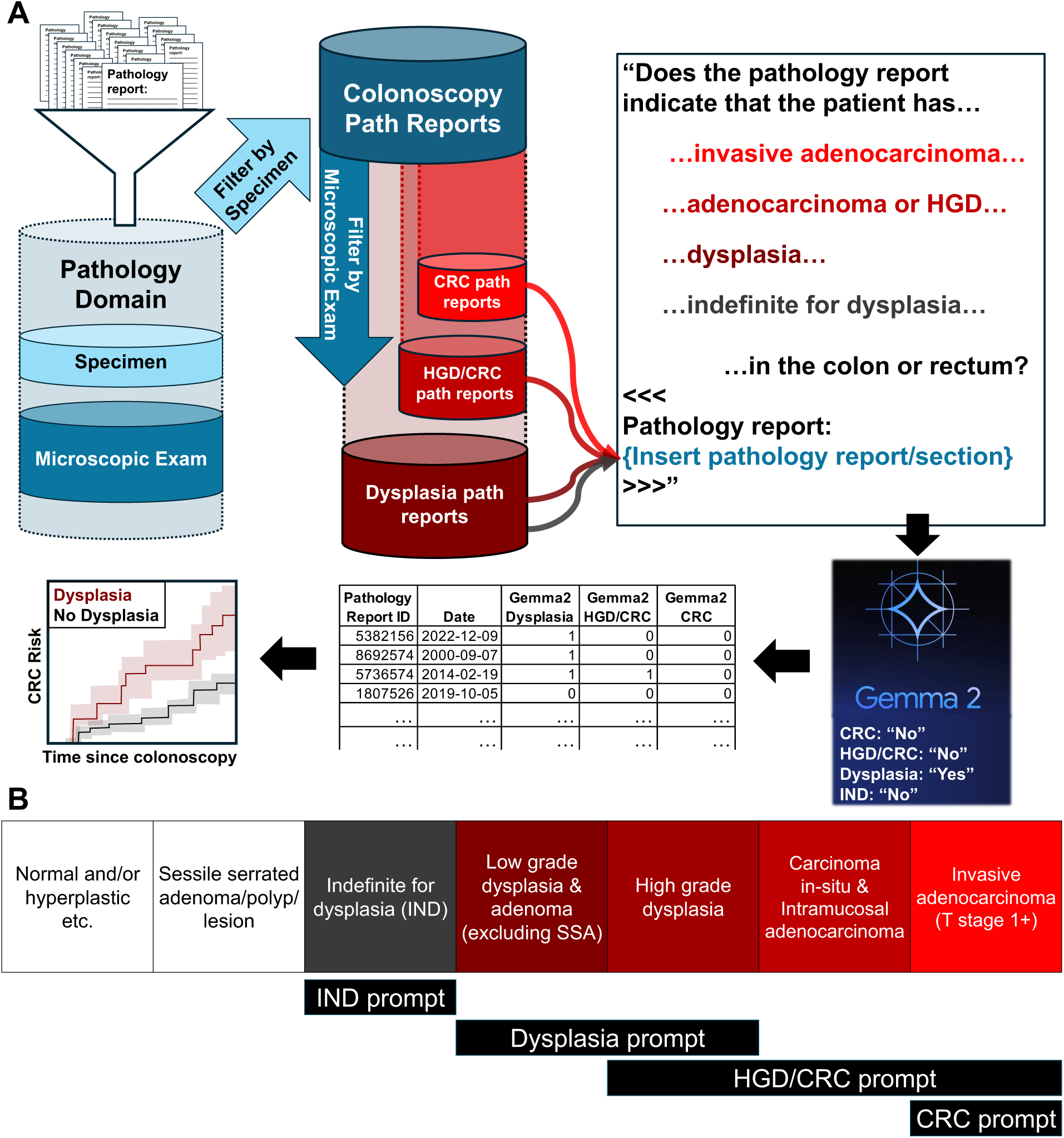
Workflow for extracting diagnoses from electronic health data. **A:** The Pathology Domain sections were used for simple term filtering designed to reduce the total number of reports fed to the LLM, while still capturing all pathology reports with possible diagnoses. Then the Microscopic Exam section of the report (or the full pathology report) is integrated into a prompt fed to the LLMs. A partial prompt is shown in the figure for illustrative purposes, and the full prompts are available in Materials and Methods and Supplementary Information. The LLM answers (“Yes” or “No”) are converted to structured data, which can be used for downstream applications, such as estimating a stratified risk of CRC based on colonoscopy findings. **B:** Diagnoses to be obtained by 4 prompts utilized in this study. By applying each of the four prompts, the most advanced diagnosis can be stratified into each of the following five buckets: no dysplasia or adenocarcinoma, indefinite for dysplasia, low-grade dysplasia, high-grade dysplasia OR intramucosal adenocarcinoma OR carcinoma in-situ, and invasive adenocarcinoma. HGD/CRC = High-grade dysplasia or adenocarcinoma; CRC = Colorectal cancer (invasive colorectal adenocarcinoma); Path = Pathology.

### Datasets and compute environment

#### Patient databases

We applied our methods to data from the nationwide Veterans Health Administration (VHA), one of the largest integrated health systems in the US. The Corporate Data Warehouse (CDW) in the Veterans Affairs (VA) contains all electronic health record (EHR) data from Veteran healthcare encounters, including notes, International Classification of Diseases (ICD) codes, and other registries such as the National Death Index (NDI) that can all be used and is intended for research purposes. In total, our provisioned CDW database contains the EHRs from 15.2 million current and former patients cared for through the VHA. This consists of roughly 6.2 billion notes, with a mean of more than 400 notes per patient. The earliest notes and other data relevant for our purposes, such as ICD codes, date back to around the year 2000, when records began to be consistently stored digitally.

The Million Veteran Program (MVP) is a research initiative where Veterans volunteer to have additional health, survey, and complete genetic data collected and made available for research in an anonymized way. To date, over one million Veterans have volunteered to become a part of this initiative. Our provisioned dataset contains 913,318 patients (version 22 data release, 8/2023). Veteran volunteers in MVP are demographically similar to patients in CDW and are a representative subsample^20^, though re-identification or any linking of clinical data is strictly disallowed to protect the privacy of MVP participants. Therefore, cohort building must be done in each dataset separately. Further, any overlap of patients and/or notes between MVP and CDW datasets is due to random chance and is not known to researchers.

#### VINCI workspaces

The VA Informatics and Computing Infrastructure (VINCI) is a platform where researchers can access clinical data from both CDW and MVP. VINCI allows researchers to analyze these data sources using various computational resources in a secured environment. Within VINCI, structured and unstructured free text data are organized in SQL tables. R and Rstudio were used to process text from SQL tables into “.txt” files for reading into compiled C++ software, which was a fork we created of the open source llama.cpp GitHub project^21^. We first used the standard 4-core CPUs available on VINCI development workspaces. We were also then able to compare results using 2 Nvidia A40 GPUs provisioned through VINCI and calculated the inference speed increase that GPU use enabled.

#### VA Pathology Domain

The VINCI team has created a dataset domain called the Pathology Domain, which takes full pathology reports and extracts certain sections based on their appropriate section headers. The resulting table contains columns representing each section header, with the associated text for each note. Our method uses the columns “Specimen”, to determine if the report described tissue from the colon or rectum, and “Microscopic Exam”, to extract the diagnosis. We also tested the method on full text of the corresponding pathology report, where available.

The Pathology domain is available in CDW to approved VINCI researchers. In MVP, the Pathology Domain data must be requested. We requested all Microscopic Exam and Specimen sections where Specimen had one of the following terms: “rectum”, “colorectal”, “rectal”, “cecum”, “colon”, “hepatic flexure”, “ileocecal valve”, “rectosigmoid”, “splenic flexure”, or “colonic flexure”. This term search was not case-sensitive and used word boundaries to identify terms.

### LLM approach and application

#### Large Language Models

The main model used is Gemma-2-9B-It-SPPO^22,23^, referred to herein as Gemma-2 (9 billion parameters). We also applied Llama-3-8B-Instruct^24^, referred to herein as Llama-3 (8 billion parameters), to all tasks/cohorts. All models used have licenses that allow commercial and research use, as required by VA policy. All models were run as “.gguf” files, the all-in-one file format used by llama.cpp^21^. For details, see Supplementary Information section “*Large Language Model (LLM) selection*”.

#### Identifying colonoscopy pathology reports in the Pathology Domain

As mentioned in *VA Pathology Domain*, the partitioned data in the MVP pathology domain was filtered to only include those where the Specimen matches colon or rectum terms. In CDW, we apply the same terms matching: “rectum”, “colorectal”, “rectal”, “cecum”, “colon”, “hepatic flexure”, “ileocecal valve”, “rectosigmoid”, or “splenic flexure”. As in MVP, this term search is not case-sensitive. This led to a corpus of relevant notes for our study totaling n=2,899,321 reports from 1,834,930 unique patients in CDW and n=279,964 reports from 170,806 unique patients in MVP. Similar filtering in Pathology Domain was previously used to validate method for extracting adenoma detection rates^25^. Pathology reports with inaccurate specimen extraction (due to typos for example) were not included in our corpus of relevant notes.

#### Tasks

Our approach identified the presence or absence of the following three concepts (clinical conditions) on any colonoscopy-associated pathology reports: any dysplasia, high-grade dysplasia and/or any adenocarcinoma (HGD/CRC), and invasive adenocarcinoma (CRC). Additionally, for the IBD-specific population, we considered a concept for indefinite for dysplasia (IND). Our definitions for each of these four concepts are as follows:

**Any confirmed dysplasia:** Presence of any dysplasia in the colon or rectum explicitly stated in the report (e.g., low-grade, mild, moderate, high-grade etc.). This includes presence of any adenoma or adenomatous lesions in the colon or rectum, excluding sessile serrated adenoma unless there is an explicit statement of sessile serrated adenoma with dysplasia. Excludes “indefinite for dysplasia” and other uncertain phrases. Adenomas were counted in definition of any dysplasia because all adenomas contain at least low-grade dysplasia (LGD).

**High-grade dysplasia and/or colorectal cancer (HGD/CRC):** Presence of HGD or any adenocarcinoma in the colon or rectum. Includes carcinoma in situ, adenocarcinoma in-situ and intramucosal adenocarcinoma. Excludes uncertain phrases such as “bordering on high-grade dysplasia”. This concept was chosen for its relevance in IBD (advanced neoplasia as a clinical outcome in treatment decision-making).

**Invasive colorectal cancer (CRC):** Presence of invasive adenocarcinoma of the colon or rectum. Invasive is defined as T stage of 1 or greater, or equivalent language (e.g., “invades into submucosa”). Excludes metastatic adenocarcinoma suspected or known to be from a different primary location (i.e. primary is not colon or rectum). Excludes uncertain phrases such as “cannot rule out invasive adenocarcinoma” and “suspicious for invasion” (further details in Supplementary Methods section *Model validation details)*.

**Indefinite for dysplasia (IND)**^26^: In IBD, presence of changes in the colon or rectum that raise concern for dysplasia but lack definitive features to confirm. This includes findings described as ambiguous due to factors such as inflammation, regenerative changes, or poor quality. Excludes explicit statements confirming the presence of dysplasia or adenomas.

We chose to evaluate these tasks as examples for our study, but the robust LLM framework described below can also distinguish other groupings if desired (such as LGD separately). However, we note that applying these tasks simultaneously does allow us to automatically classify the most advanced lesion in each pathology report (Figure 1B). For example, if the report is positive for Dysplasia and negative for HGD/CRC, we can conclude that the sample has LGD. Similarly, if the report is positive for HGD/CRC and negative for CRC, we can conclude that the sample has HGD or intramucosal CRC only. While both HGD/CRC and CRC are indications for resection in IBD, prognosis for these may differ in the non-IBD population. Furthermore, distinguishing between invasive and noninvasive adenocarcinoma or HGD is useful for incidence reporting and epidemiological studies, so we separated these herein.

#### Creation of plausible sets of notes

We then used simple search terms to reduce the number of colonoscopy pathology reports to only include those potentially diagnostic of a particular concept. For CRC, these were reports where the Microscopic Exam section text matched “%carcinoma%”, “%tumor%”, or “%invasi%”, where “%” represents a wildcard. The pathology reports matching this search are considered a part of the “plausible set” for CRC identification. The plausible sets for dysplasia and HGD/CRC were expanded to include more search terms identifying those diagnoses. See Supplementary sections “*HGD/CRC ascertainment*”, “*Dysplasia ascertainment*”, and “*Indefinite for dysplasia ascertainment*” for details. Supplementary Table S1 contains the exact numbers of reports considered for all patient cohorts at each step of filtering across tasks, starting with 16.3 million and 2.6 million total pathology reports in CDW and MVP, respectively.

#### LLM prompt development

LLMs require a ‘prompt’ to perform a given task. A ‘prompt’ is defined as the input text given to the model. The model then evaluates the prompt and generates additional text. The prompt we provide to the model consists of some text that defines the task and the question to be answered. Additionally, the prompt includes the text from the pathology report or section to be evaluated. We developed the prompt using 48 pathology report Microscopic Exam sections where the Specimen section matched colon or rectum terms (Figure 2). For details on the evolution of the prompts, see Supplementary Methods section “*Lessons learned from applying LLMs in structured and unstructured pathology report data*”. No a priori performance targets were applied after this iteration. Then for each task, the final validation sets to evaluate the LLMs excluded all reports previously chart reviewed that were considered part of the development sets for a given task.

**Figure 2:**
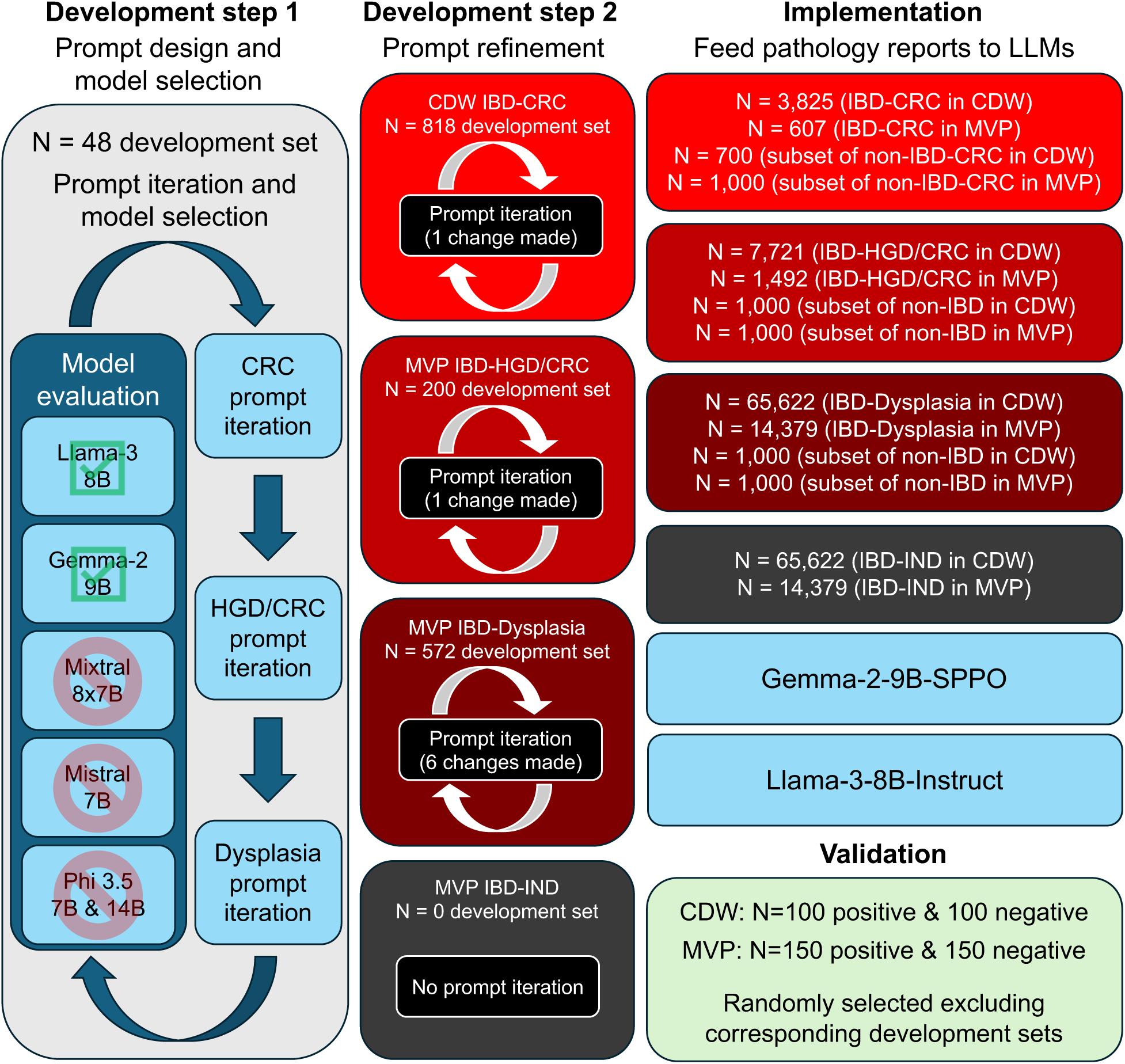
Workflow of large language model selection and prompt development. Step 1 consisted of a small set of manually de-identified reports which allowed for rapid prompt iteration and evaluation of multiple models. Once models were selected and uploaded to VINCI, further prompt iteration was performed until the errors could no longer be obviously fixed by an improved prompt. Note that indefinite for dysplasia prompt did not have a development set or any prompt iteration, nor was it used in the process of model selection. Finally, the LLMs were used for implementation and validation. A subset was used for non-IBD reports, and the size of the subset was estimated based on model prevalence in the IBD cohort to ensure enough model positives and negatives to allow for stratified sampling.

#### Determining presence vs. absence of given diagnosis using LLM

For each report in the plausible set of pathology reports, we feed either the Microscopic Exam section (or the full text pathology note) to an LLM which determines if an individual has a specific pathological diagnosis (Figure 1A). The input text is integrated into the prompt (see Supplementary Methods sections “*CRC ascertainment*”, “*HGD/CRC ascertainment*”, “*Dysplasia* ascertainment” and “*Indefinite for dysplasia ascertainment*” for exact prompts used). The model then responds “Yes” or “No”. This response is recorded as an output “.txt” file with the corresponding ID of the pathology domain entry. Llama.cpp^21^ is used for model inference.

#### Model validation

Validation was performed independently in IBD and non-IBD populations using the same models and three prompts (see Supplementary Methods for cohort creation). For each validation set, either N=100 (CDW) or N=150 (MVP) randomly selected putative positive cases and the same number of putative negative cases were selected for review. Putative positive (negative) cases were defined as cases where Llama-3 responded “Yes” (“No”). Validation was performed by two independent, blinded reviewers (BJ & AD). Disagreements between AD and BJ, of which there were 160 across 4,007 total reviewed pathology reports (< 4% of reviewed pathology reports), were resolved by a third, blinded reviewer (HE). If HE lacked certainty, which occurred for 4 reports total, LJJ resolved the disagreement as a blinded 4th reviewer. Cohen’s kappa was used to measure inter-reviewer agreement between BJ and AD. Validation was done at the level of the pathology report, consistent with the LLM prompt asking if the given features are present in any colon or rectal sample. Validation was performed independently for each of the tasks, even if notes overlapped by chance in validation sets across tasks.

We performed validation only in the “plausible set” of notes that passed our search term filters (Supplementary Table S1) and recorded run-times (Supplementary Table S2). Considering the very low expected prevalence (potentially zero) outside of these filters, this approach provides a more informative assessment of the LLMs’ performance, as we are considerably more likely to include some false negative cases in our validation (see Supplementary methods for details). For testing generalizability and validity, we also evaluated performance using full pathology reports as LLM prompt input. Full-text pathology reports were considered in validation analyses only and were not used in prompt development, which only used semi-structured reports.

#### Performance metrics

We provide an estimate of the prevalence of cases in the reports from the plausible set, as well as the positive predictive value (PPV), negative predictive value (NPV), sensitivity (recall), specificity, F1-score, and Matthew’s Correlation Coefficient (MCC). In order to better compare across tasks with varying class ratios (prevalence), we also show the calibrated precision (PPVc) and calibrated F1 score (F1c) based off of previous work by Siblini et al.^19^, computed using a reference ratio *π*_0_=0.5. These calibrated metrics correct for the dependence of precision-based performance metrics on the positive class ratio. More specifically, if true positives are rare, precision will be lower than if the same model were evaluated on a more balanced dataset. For a given reference ratio, *π*_0_, the calibrated metrics provide the expected PPV and F1-score for a dataset with a positive class ratio of *π*_0_^19^.

Because we use stratified sampling, i.e., selecting N model-predicted positive and N model-predicted negative reports for validation, calculating the performance metrics listed above requires corrections to account for the conditional probabilities introduced by our sampling. The explicit forms of these equations are derived in Supplementary Methods section “*Calculating performance metrics”*. This approach helps minimize the number of cases needed for validation, especially when prevalence is imbalanced, and builds on previous work by Liu et al.^28^. Figure 2 shows the flowchart for all steps of LLM development through validation, including numbers of all pathology reports evaluated for each task.

## Results

We applied LLMs to extract pathologic diagnoses from text in the VA Pathology Domain (Figure 1) and free-text pathology reports. We tested two LLMs (Gemma-2 and Llama-3) for each classification task. After prompt development, we validated our methods by comparing model predictions to blinded chart review by two independent reviewers evaluating randomly chosen sets of reports for each task (any dysplasia, HGD/CRC and CRC) in each patient cohort (IBD and non-IBD) and dataset (MVP and CDW). The overall agreement between the two reviewers was excellent for these tasks, with Cohen’s kappa ranging from 89-97%. The task of indefinite for dysplasia (IND) was also evaluated in the IBD cohort.

### Large language models extract pathologic diagnoses with high accuracy in patients with IBD

In model validation using strictly distinct reports from those used for prompt development (see Materials and Methods), all tasks in IBD achieved excellent performance using LLM Gemma-2 (Table 1). Metrics such as precision (PPV) and F1 are dependent on the class ratio, meaning that the uncalibrated F1 and PPV are not as useful for comparing across tasks with very different prevalences^19^. This is especially relevant when comparing the main 3 diagnostic tasks to the IND task, which is relatively rare in the plausible set of pathology reports filtered using the dysplasia terms in IBD. The calibrated F1 score, which combines calibrated precision and recall, for IND was 98.6% (95% CI 96.9-99.8%) in MVP and 95.6% (95% CI 92.9-98.9%) in CDW when using the Microscopic Exam section as input. In comparison, the task with the highest calibrated F1 score was diagnosis of CRC in MVP data (F1c = 99.3% (95% CI 98.7-99.8%)).

**Table 1:** Validated performance results for IBD patients in MVP and CDW using Gemma-2. Shading progression has lower value (red) = 0.5, middle value (white) = 0.9 and upper value (green) = 1. Invasive CRC = Invasive colorectal cancer (invasive colorectal adenocarcinoma); HGD/CRC = High-grade dysplasia and/or colorectal adenocarcinoma; IND = indefinite for dysplasia; IBD = Inflammatory bowel disease; PPV = Positive predictive value; NPV = Negative predictive value; LB = Lower bound; UB = Upper bound; PPVc = Calibrated positive predictive value (calibrated precision)^19^; F1c = Calibrated F1 score^19^; MCC = Matthew’s correlation coefficient. 95% confidence intervals for PPV and NPV were approximated using the binomial distribution (see Supplementary Methods).

| Task | Source | Model prevalence estimate | PPV (LB - UB) | NPV (LB - UB) | PPV<sub>c</sub> | Recall (Sensitivity) | Specificity | F1 | F1<sub>c</sub> | MCC | Cohen's kappa |
| --- | --- | --- | --- | --- | --- | --- | --- | --- | --- | --- | --- |
| CRC | MVP | 0.257 | 0.962 (0.92 - 0.99) | 1.000 (0.98 - 1.00) | 0.986 | 1.000 | 0.987 | 0.980 | 0.993 | 0.974 | 0.889 |
| CRC | CDW | 0.266 | 0.916 (0.84 - 0.99) | 0.967 (0.92 - 0.99) | 0.971 | 0.910 | 0.969 | 0.913 | 0.939 | 0.881 | 0.910 |
| HGD/CRC | MVP | 0.176 | 0.921 (0.84 - 0.99) | 0.990 (0.96 - 1.00) | 0.983 | 0.953 | 0.983 | 0.937 | 0.968 | 0.924 | 0.947 |
| HGD/CRC | CDW | 0.219 | 0.941 (0.82 - 1.00) | 0.990 (0.95 - 1.00) | 0.983 | 0.962 | 0.983 | 0.951 | 0.973 | 0.938 | 0.970 |
| Dysplasia | MVP | 0.228 | 0.947 (0.90 - 0.99) | 0.993 (0.96 - 1.00) | 0.984 | 0.976 | 0.984 | 0.961 | 0.980 | 0.950 | 0.920 |
| Dysplasia | CDW | 0.313 | 0.964 (0.90 - 1.00) | 1.000 (0.96 - 1.00) | 0.983 | 1.000 | 0.984 | 0.982 | 0.992 | 0.974 | 0.950 |
| IND | MVP | 0.030 | 0.752 (0.57 - 0.93) | 1.000 (0.98 - 1.00) | 0.990 | 0.983 | 0.992 | 0.852 | 0.986 | 0.856 | 0.931 |
| IND | CDW | 0.031 | 0.526 (0.42 - 0.82) | 0.999 (0.97 - 1.00) | 0.974 | 0.938 | 0.985 | 0.674 | 0.956 | 0.696 | 0.781 |

We found slightly lower performance when using LLM Llama-3 (Supplementary Table S3). As expected, smaller LLMs with fewer parameters were less accurate in the 4 tasks for IBD (Supplementary Table S4). GPU use significantly reduced run times (Supplementary Table S2).

### Validation of large language model approach in non-IBD colorectal dysplasia and cancer

We then applied the same validation approach, with no changes to model prompts, to records from patients without IBD (i.e., no IBD colitis ICD code found in patient clinical history) and again achieved highly accurate results in the 3 relevant tasks for non-IBD reports (Table 2). Specifically, we found that the F1-score for identifying dysplasia in patients without IBD was ∼99% using both Gemma-2 and Llama-3 (Table 2, Supplementary Table S3). F1-scores were slightly lower but still excellent for HGD/CRC (>96%) and CRC (>95%) using Gemma-2. These results highlight the flexibility of the LLM approach for similar histopathologic diagnoses in different patient groups.

**Table 2:** Validation performance results for non-IBD colitis patients in MVP and CDW using Gemma-2. Shading progression has lower value (red) = 0.5, middle value (white) = 0.9 and upper value (green) = 1. CRC = invasive colorectal cancer (invasive colorectal adenocarcinoma); HGD/CRC = High-grade dysplasia and/or colorectal adenocarcinoma; IND = indefinite for dysplasia; IBD = Inflammatory bowel disease; PPV = Positive predictive value; NPV = Negative predictive value; LB = Lower bound; UB = Upper bound; PPVc = Calibrated positive predictive value (calibrated precision)^19^; F1c = Calibrated F1 score^19^; MCC = Matthew’s correlation coefficient. 95% confidence intervals for PPV and NPV were approximated using the binomial distribution (see Supplementary Methods).

| Task | Source | Model prevalence estimate | PPV (LB - UB) | NPV (LB - UB) | PPV<sub>c</sub> | Recall (Sensitivity) | Specificity | F1 | F1<sub>c</sub> | MCC | Cohen's kappa |
| --- | --- | --- | --- | --- | --- | --- | --- | --- | --- | --- | --- |
| CRC | MVP | 0.522 | 0.982 (0.94 - 1.00) | 0.908 (0.85 - 0.95) | 0.982 | 0.921 | 0.978 | 0.950 | 0.950 | 0.895 | 0.920 |
| CRC | CDW | 0.525 | 0.971 (0.92 - 1.00) | 0.951 (0.88 - 0.99) | 0.970 | 0.957 | 0.968 | 0.964 | 0.963 | 0.924 | 0.940 |
| HGD/CRC | MVP | 0.306 | 0.977 (0.93 - 1.00) | 0.979 (0.94 - 1.00) | 0.990 | 0.953 | 0.990 | 0.965 | 0.971 | 0.949 | 0.947 |
| HGD/CRC | CDW | 0.411 | 0.966 (0.90 - 1.00) | 0.990 (0.94 - 1.00) | 0.976 | 0.985 | 0.976 | 0.975 | 0.981 | 0.958 | 0.950 |
| Dysplasia | MVP | 0.896 | 0.986 (0.95 - 1.00) | 0.993 (0.96 - 1.00) | 0.889 | 0.999 | 0.890 | 0.992 | 0.941 | 0.933 | 0.953 |
| Dysplasia | CDW | 0.870 | 0.988 (0.95 - 1.00) | 0.990 (0.94 - 1.00) | 0.923 | 0.998 | 0.923 | 0.993 | 0.959 | 0.949 | 0.920 |

### Accuracy of applying LLM methods to full text pathology report

To evaluate the generalizability of our model to environments that do not contain semi-structured resources such as the VA Pathology Domain, we applied our LLM approach to the full pathology report to evaluate performance and found excellent measures using Gemma-2 (Table 3). In IBD, the task with the best performance was presence of any dysplasia (F1 = 96.6% (95% CI 92.0-100%)) and lowest performance of the 3 main tasks was diagnosis of CRC (F1 = 88.1% (95% CI 82.0-93.8%)). Higher values were found for the calibrated F1-scores. While both Gemma-2 and Llama-3 were trained with context lengths up to 8,192 tokens, and the full notes never exceeded these thresholds, performance decreases slightly when using Llama-3 (Supplementary Table S3). We also found similar performance results to Gemma-2 alone when requiring either or both models to answer ‘Yes’ for a report to be deemed a positive case (Supplementary Table S5).

**Table 3:**
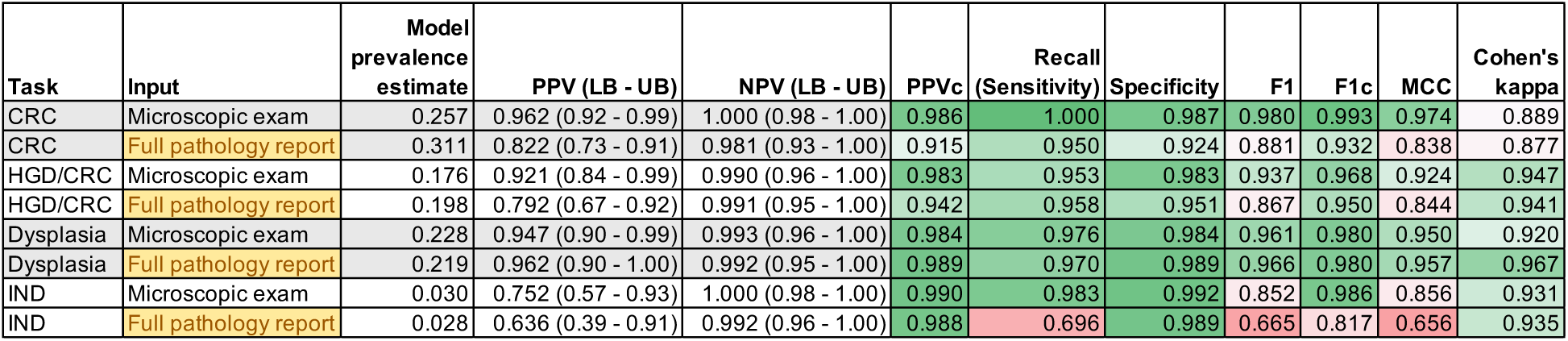
Validation results using full pathology report in IBD population in MVP. Comparison of Microscopic Exam section (these rows are repeated from Table 1) and full pathology report as input to LLM. Full pathology reports evaluated by LLMs in all cases where the Pathology Domain entry had a matching full note. There were instances where the full note was not available, and so the number validated for the full pathology report was less than the total number validated for “Microscopic exam”. All analyses still had >100 model positive and model negative cases validated. See Supplementary Table S6 for details on pathology report numbers. 95% confidence intervals for PPV and NPV were approximated using the binomial distribution (see Supplementary Methods). CRC = invasive colorectal cancer (invasive colorectal adenocarcinoma); HGD/CRC = High-grade dysplasia and/or colorectal adenocarcinoma; IND = indefinite for dysplasia; IBD = Inflammatory bowel disease; PPV = Positive predictive value; NPV = Negative predictive value; LB = Lower bound; UB = Upper bound; PPVc = Calibrated positive predictive value (calibrated precision)^19^; F1c = Calibrated F1 score^19^; MCC = Matthew’s correlation coefficient.

| Task | Input | Model prevalence estimate | PPV (LB - UB) | NPV (LB - UB) | PPV<sub>c</sub> | Recall (Sensitivity) | Specificity | F1 | F1<sub>c</sub> | MCC | Cohen's kappa |
| --- | --- | --- | --- | --- | --- | --- | --- | --- | --- | --- | --- |
| CRC | Microscopic exam | 0.257 | 0.962 (0.92 - 0.99) | 1.000 (0.98 - 1.00) | 0.986 | 1.000 | 0.987 | 0.980 | 0.993 | 0.974 | 0.889 |
| CRC | Full pathology report | 0.311 | 0.822 (0.73 - 0.91) | 0.981 (0.93 - 1.00) | 0.915 | 0.950 | 0.924 | 0.881 | 0.932 | 0.838 | 0.877 |
| HGD/CRC | Microscopic exam | 0.176 | 0.921 (0.84 - 0.99) | 0.990 (0.96 - 1.00) | 0.983 | 0.953 | 0.983 | 0.937 | 0.968 | 0.924 | 0.947 |
| HGD/CRC | Full pathology report | 0.198 | 0.792 (0.67 - 0.92) | 0.991 (0.95 - 1.00) | 0.942 | 0.958 | 0.951 | 0.867 | 0.950 | 0.844 | 0.941 |
| Dysplasia | Microscopic exam | 0.228 | 0.947 (0.90 - 0.99) | 0.993 (0.96 - 1.00) | 0.984 | 0.976 | 0.984 | 0.961 | 0.980 | 0.950 | 0.920 |
| Dysplasia | Full pathology report | 0.219 | 0.962 (0.90 - 1.00) | 0.992 (0.95 - 1.00) | 0.989 | 0.970 | 0.989 | 0.966 | 0.980 | 0.957 | 0.967 |
| IND | Microscopic exam | 0.030 | 0.752 (0.57 - 0.93) | 1.000 (0.98 - 1.00) | 0.990 | 0.983 | 0.992 | 0.852 | 0.986 | 0.856 | 0.931 |
| IND | Full pathology report | 0.028 | 0.636 (0.39 - 0.91) | 0.992 (0.96 - 1.00) | 0.988 | 0.696 | 0.989 | 0.665 | 0.817 | 0.656 | 0.935 |

## Discussion

We have shown that LLMs are powerful, potentially generalizable tools for accurately extracting important information from clinical semi-structured and unstructured text and which require little human-led development. With validated performance by blinded manual chart review also shown in Million Veteran Program data, we expect this approach will enable large-scale health research studies that can incorporate patient genomics in disease risk assessment and prediction. Another strength of this work is that the methods are relatively simple. While not explicitly tested, we expect our findings to adapt relatively easily to other pathological diagnoses, healthcare systems, patient populations, and time periods. No aspect of the prompt or models used were specific to the VA or our cohorts, and no additional model fine-tuning was performed. The barriers to implementation are minimal; any researcher can download the llama.cpp^21^ GitHub, add their desired prompt, compile, and begin development. Our repository is available on GitHub for the community.

While previous NLP approaches show excellent performance in identifying common features like adenoma^10,11,14^, few have maintained excellent performance with thorough testing of their approaches to identify rarer advanced features such as HGD, carcinoma in situ, and invasive adenocarcinoma. Additionally, few have tested approaches in differing contexts, such as differing geographical locations, practice types (e.g., academic vs. private practice), and compensation structures (e.g., salary vs. fee-for-service)^16^. When attempted across 4 practice sites, Carrell et al. report an F1-score of 95% for identifying adenoma and highlight the considerable time-consuming challenges they encountered in adapting the NLP system^16^. The most comparable analysis in our study to previously published algorithms was the task of identifying any dysplasia in the non-IBD colitis cohort, where Gemma-2 had an F1-score of 99.2% (95% CI 98.2-100%) and Llama-3 had an F1-score of 99.2% (95% CI 98.1-100%) in MVP, and even higher scores were found in CDW. Published analyses in similar cohorts have similarly high F1-scores, such as Bae et al., who report an F1-score of 99% for identifying the presence of “conventional adenoma”^14^, and Nayor et al., who report a perfect F1-score of 100% for identifying “adenoma”^11^. Supplementary Table S7 shows a comparison of performance using previous methods across comparable tasks. Notably, code for these rule-based approaches was not made available in most cases to enable direct application in our data.

While LLMs remain computationally expensive, the size and associated compute cost of proficient models has reduced drastically, with the best small (9 billion parameter), open-weight model available at the time of this work (Gemma-2-9b-it^22^, released June 27, 2024) generally performing better than the largest proprietary models from a year prior (GPT-4-0613^29^, released June 13, 2023)^30^. If such improvements in efficiency continue, boosted by potential advancements in the underlying transformer architecture^31^, LLMs will become more attractive in domains where the current computational expense makes their use unfeasible. Importantly, the LLM approach is robust and can be readily adapted with minimal, if any, tweaks to prompts/code as these newer models become available online. Even without further improvements in the models themselves, the increasing availability of GPUs and the throughput of new chip architectures^32–34^ may make current models a viable alternative to data structuring at scale. Due to the ease of our model implementation, with results as accurate as more complicated rule-based approaches, we suggest an LLM approach for many free-text classification tasks in biomedical research going forward.

Our work has some limitations. First, while we expect our approach to adapt more flexibly to different settings, we did not explicitly test our LLM approach in other large-scale EHR datasets beyond the VHA, though recent success applying LLMs in other health systems has been shown^17,18^. Second, without long-term access to Graphics Processing Units (GPUs), we could not feasibly test larger models which may overcome some of the shortcomings seen in smaller models; this addition can be expected to increase performance above what we find herein. Finally, we could not rule out overlap between MVP and CDW reports, though our results in either cohort considered alone are sufficient validation compared to previously published work.

Ongoing work includes adapting our approach to detect stage and location of cancers, identifying features of dysplasia (size, shape, type, location, inflammation level, etc.) and endoscopic resection details, as well as incorporation with genetic data. Some tasks, such as identifying IBD sub-types and dates of diagnosis, may require larger models that are more capable of handling longer input text. Nonetheless, the general framework lends itself to many applications beyond the use cases analyzed here, including the potential for real-time data integration in models used to aid in shared decision-making (so-called medical digital twins)^35^. While we show that the applications for research are immediate (low compute requirements, free access to open weight models that do not risk patient privacy), these exciting opportunities for LLM integration into existing healthcare systems will have to overcome certain challenges for real-world deployment (e.g., IT infrastructure, potential biases, and user trust) that will be the focus of many future quality improvement studies.

Accurate clinical data is essential for understanding trends in patient disease risk and for predictive models to be clinically useful. In an era of increasing opportunities for personalized medicine, we show that large language models offer a very useful tool for quickly and accurately obtaining relevant patient data to potentially inform medical decisions in real time.

## Code availability

Llama.cpp fork will be made available at https://github.com/bdj34/llama.cpp_data_extraction, which includes the main inference code from llama.cpp. Our specific implementation can be found at https://github.com/bdj34/llama.cpp_data_extraction/tree/brian-features/examples/data-extraction which includes all prompts and custom parameters required to reproduce our work. Large language models, in gguf format, used in this work are stored at https://huggingface.co/briandj97/models_used.

## Data availability

A CSV (all_results.csv) with results from all validated runs, including additional details such as prevalence of model positives, exact numbers validated, and the full confusion matrix, will be available in the supplementary information. This CSV is the source for all three main text tables and Supplementary Tables S3-S5. Raw data access is reserved for VA investigators with appropriate research approvals.

## Author contributions

Study conception and design: KC, BJ

Data collection and curation: BJ, TB, XH, ML, AE, HE, LJJ

Model development and formal analyses: BJ, KC, SG, SCS

Paper writing: BJ, KC, SG, SCS

## Data availability

A CSV (all_results.csv) with results from all validated runs, including additional details such as prevalence of model positives, exact numbers validated, and the full confusion matrix, will be made available in the supplementary. This CSV is the source for all three main text tables and Supplementary Tables S3-S5. Raw data access is reserved for VA investigators with appropriate research approvals.

## Code availability

Llama.cpp fork and our specific implementation will be made available on Github, which includes all prompts and custom parameters required to reproduce our work. Large language models, in gguf format, are stored and publicly available at Hugging Face.

## Data Availability

All processed data produced in the present work are contained in the manuscript.
Raw data access is reserved for VA investigators with appropriate research approvals.

## Acknowledgements

This research is based on data from the Million Veteran Program, Office of Research and Development, Veterans Health Administration, and was supported by MVP070 as well as Merit Review Award I01 BX005958 from the United States (U.S.) Department of Veterans Affairs Biomedical Laboratory Research and Development Service. The contents do not represent the views of the U.S. Department of Veterans Affairs or the United States Government. This work was supported by AGA Research Foundation (AGA Research Scholar Award AGA2022-13-05), NIH grants (R01 CA270235, P30 CA023100), and National Library of Medicine Training Grant (NIH grant T15LM011271). The study was supported in part by the NIDDK-funded San Diego Digestive Diseases Research Center (P30 DK120515).

