## Supplementary Information for "Large language models for extracting histopathologic diagnoses of colorectal cancer and dysplasia from electronic health records"

### Supplementary Methods

***Potential Million Veteran Program (MVP) and Corporate Data Warehouse (CDW) overlap***

We considered MVP and CDW as separate cohort building tasks because re-identification of MVP participants is strictly disallowed. We can conservatively estimate the fraction of pathology reports that overlap for the case with most likely overlap (rarest), colorectal cancer within inflammatory bowel disease (IBD-CRC). In MVP, the IBD-CRC plausible set consists of 607 distinct pathology reports. In CDW, the IBD-CRC plausible set consists of 3,825 distinct pathology reports. If we assume the 607 are a subset of the 3,825, we can estimate the fraction of the notes that overlap. In this case, we draw 307 pathology reports to review in MVP. However, 159 of these are putative positive reports by either Llama-3 or Gemma-2 and 148 are negative by both. Overlaps are more likely in the relatively less common positive reports, so we consider these only. Now, we know that 159 represents all the positive cases by either model from the IBD-CRC plausible set in MVP. There are roughly between 1,003 and 1,100 putative positives of the 3,825 reports in the IBD-CRC plausible set. Llama-3 was positive for 1,003 of 3,825 reports, but Gemma-2 was only run on a random subset of 700 of the plausible set reports. Let's assume 1,000 reports are positive in CDW, rounding down slightly which will increase our estimate of overlaps. As we randomly select 100 from the CDW positives, our expected number of overlapping notes is 100*(159/1000) = 15.9. Therefore, we expect that < 16% of our IBD-CRC putative positive reports overlap. In the putative negative reports, the larger cohort of non-IBD colitis, and/or the larger plausible sets such as those for dysplasia and HGD/CRC, we expect a much smaller fraction of the reports to overlap.

Using the mean of the hypergeometric distribution, n*K/N, where n is the number of draws (100 for validated CDW putative positives), K is the number of successes (150 for validated MVP putative positives) and N is the population size (the prevalence of model positives multiplied by the plausible set notes in CDW), we can estimate the expected overlap for any matching cohort and task. For example, in non-IBD CRC this calculation would be roughly 100*(150/(104708*0.5)) = 0.287, so < 1% of positive and negative reports would be expected to overlap. For IBD dysplasia, we would expect roughly 100*(150/(65622*0.3)) = 0.762 and again < 1% of reports would be expected to overlap.

While CDW is a much larger dataset, the MVP dataset includes genotype data, which can be used for more analyses such as development of polygenic risk scores (PRS). As such, it was useful to build cohorts and validate our methods in both data sources. While there are slight differences in the partitioning and organization of the data, as well as the potential for chance overlap across tasks, it is nonetheless reassuring to see similar results across MVP and CDW.

***VINCI workspace specifications***

Our VINCI development workspace CPUs were the standard 4-core, 2.3 GHz "Intel Core Processor (Haswell, no TSX)". The Haswell architecture was first introduced in 2013. Upon request, these workspaces were allocated additional memory, with one having 32GB RAM and the other having 20GB.

We also used a VINCI Linux development workspace with 2 Nvidia A40 GPUs for running the “indefinite for dysplasia” diagnostic task to compare with CPU runtimes and show the running time differences (CPU vs GPU) in Supplementary Table S2.

***Large Language Model (LLM) selection***

Over the course of this study, new large language models were continuously released. If it was clear that the new models were better than the ones we were currently using, we replaced them until August 1, 2024. We tested models on a set of 48 challenging pathology report Microscopic Exam sections, as detailed in Methods section “*LLM prompt development”*. These 48 reports were selected because they presented unique challenges, including reports with adenocarcinoma in different organs and those which conveyed uncertainty in the diagnosis. Many of the better models were able to score 48/48 on this challenging set.

We initially started work with two fine-tuned models based on Mistral-7B[1], Starling-LM-7B-alpha[2] and OpenHermes-2.5-Mistral-7B[3]. “Fine-tuning” in this case refers to less computationally expensive post-training, which may be performed by different individuals or organizations than those who trained the original models. We also considered the Mixtral-8x7B-Instruct[4] model, though this larger model was too expensive to feasibly run for all of our tasks and cohorts. The performance of these models was not validated as they were used only for initial exploration. Once released in April 2024, we incorporated Llama-3-8B-Instruct[5] as it became clear from external benchmarks[6,7] and our testing that it was the best performing model at that size at the time. Again, once released in June 2024, we incorporated Gemma-2-9B-SPPO, a “Self-Play Preference Optimization”[8] fine-tuned model based on Gemma2-9b-Instruct[9]. As noted, our model choices were subject to the constraints of our initially CPU-only hardware. While we tested larger models locally, such as Gemma-2-27B-Instruct[9] and Llama-3-70B-Instruct[5], it was not feasible to use these models at scale given our resource limitations.

After completing our work with Llama-3-8B and Gemma-2-9B, we decided to test the smaller versions of these models that had been released in the meantime. These include Llama-3.2-3B-Instruct[10] and Gemma-2-2B[9]. As expected, these models do not perform as well as their larger counterparts (see Supplementary Table S4).

None of the models used were trained by us and none of the fine-tuning focused specifically on clinical applications. All the models used have permissive licenses that allow commercial and research use, as required by VA policy.

As mentioned, all models were run as “.gguf” files, the all-in-one file format used by llama.cpp[11]. Gemma-2-9b-SPPO and Llama-3-8B-Instruct were run at fp16 precision and were converted from Safetensors format using llama.cpp. “Quantized” versions of Mixtral-8x7B-Instruct (Q5_K_M), Llama-3.2-3B-Instruct (Q8_0) and Gemma-2-2b-It (Q8_0) were used to fit the model within our RAM requirements or, in the case of the smaller models, to make uploading to VINCI easier. This quantization stores the model parameters at reduced precision[12]. Links to our huggingface repository with the ".gguf" file for each model, and the original source repositories, are listed below:

Our repository (all models we used in gguf format):
https://huggingface.co/briandj97/models_used
Gemma-2-9B-It-SPPO: https://huggingface.co/UCLA-AGI/Gemma-2-9B-It-SPPO-Iter3
Llama-3-8B-Instruct: https://huggingface.co/meta-llama/Meta-Llama-3-8B-Instruct
Mixtral-8x7B-Instruct (not validated): Q5_K_M quant from
https://huggingface.co/TheBloke/Mixtral-8x7B-Instruct-v0.1-GGUF/tree/main
Gemma-2-2b-It: Q8_0 quant from https://huggingface.co/bartowski/gemma-2-2b-it-GGUF
Llama-3.2-3B-Instruct: Q8_0 quant from https://huggingface.co/bartowski/Llama-3.2-3B-Instruct-GGUF

***IBD colitis and non-IBD colitis cohorts***

To split the cohort between patients with versus without IBD colitis, we use a modified version of a previously validated IBD ascertainment algorithm in VHA data[13]. Here we focused on identifying patients with IBD colitis specifically, who are at risk of colitis-associated dysplasia and colorectal cancer: ulcerative colitis, IBD-unclassified, and Crohn's colitis. Our ascertainment algorithm thus required at least 2 ICD codes matching to ones in the following list: ICD-10 codes K51.x (excluding K51.4x), K50.1x, K50.8x, K52.3, ICD-9 codes 555.1x, 555.2x, 556.x. These codes must be present on at least 2 encounters (dates) with at least one in an outpatient setting.

The non-IBD cohort comprised patients without *any* of the above listed ICD codes. Therefore, patients with only one of the above-listed ICD codes (MVP: n = 13,158; 1.44%. CDW: n = 75,925; 0.59%) were excluded from both cohorts due to their uncertain history of IBD colitis based on ICD codes.

***Microscopic description and diagnosis sections in Pathology Domain***

For the purposes of this work, we restricted our assessment to the "Microscopic Description" section, which is often used interchangeably with "Microscopic Exam". In addition to this section, there is an alternative field, in Pathology Domain called "Diagnosis". The text from "Microscopic Exam" and "Diagnosis" typically contains the same information, albeit with a different section title. This is shown by the fact that over 80% of the Pathology Domain in CDW contains only one of these sections, with the other being NULL. Roughly 98% of the Pathology Domain in CDW contains at least one of these sections (i.e. either is not NULL).

***CRC ascertainment***

The prompt given to the LLM for invasive CRC identification is as follows:

*"The text provided is a pathology report, with samples originating from the colon or rectum unless specified otherwise. We are interested in identifying whether invasive adenocarcinoma (stage greater than or equal to 1) is present in *any* colon or rectal sample. Without definite invasion identified, conditions such as 'high-grade dysplasia', 'in-situ [adeno]carcinoma', or 'intramucosal*

*[adeno]carcinoma' are not typically classified as invasive adenocarcinoma. If the sample is classified as having adenocarcinoma without further specification, this typically implies invasive adenocarcinoma. Answer yes or no to the following question, matching the format 'Answer: Yes' or 'Answer: No'. Then, explain your reasoning. Does the pathology report indicate that the patient has an invasive adenocarcinoma in any colon or rectal sample?*
<<<
*Pathology report:*
*{Insert Microscopic Exam text or full-text pathology note}*
>>>
*Does the pathology report indicate that the patient has an invasive adenocarcinoma in any colon or rectal sample?*
*Answer:"*

***HGD/CRC ascertainment***
We adapted our CRC ascertainment approach in a straightforward way to also identify high-grade dysplasia and non-invasive adenocarcinoma.

For HGD/CRC, we apply a similar regular expression filtering to the CRC algorithm with an expanded term set to capture high-grade dysplasia as well as intramucosal adenocarcinoma and carcinoma in-situ.

Invasive CRC search terms: '%carcinoma%', '%tumor%', '%invasi%'
Additional HGD/CRC search terms: '%HGD%', '%high-grade%', '%high grade%', '%in situ%', '%in-situ%', '%intramucosal%'

Similar to CRC ascertainment, the prompt for HGD/CRC identification is as follows:

*"The text provided is a pathology report, with samples originating from the colon or rectum unless specified otherwise. Answer yes or no to the following question, matching the format 'Answer: Yes' or 'Answer: No'. Then, explain your reasoning. Does the pathology report indicate that the patient has adenocarcinoma or high-grade dysplasia in any colon or rectal sample?*
<<<
*Pathology report:*
*{Insert Microscopic Exam text or full-text pathology note}*
>>>
*Does the pathology report indicate that the patient has adenocarcinoma or high-grade dysplasia in the colon or rectum?*

*Answer:"*

***Dysplasia ascertainment***
In a similar manner, we expanded the dysplasia term set as follows, including the CRC and HGD/CRC terms so that these are a subset of the Dysplasia matches.

Invasive CRC search terms: ‘%carcinoma%’, ‘%tumor%’, ‘%invasi%’
HGD/CRC search terms: ‘%HGD%’, ‘%high-grade%’, ‘%high grade%’, ‘%in situ%’, ‘%in-situ%’, ‘%intramucosal%’
Additional dysplasia search terms: ‘%dysplas%’, ‘%lgd%’, ‘%low grade%’, ‘%low-grade%’, ‘%dalm%’, ‘%adenoma%’

Similar to the other ascertainments, the prompt for dysplasia identification is as follows:

"*The text provided is a pathology report. Answer yes or no to the following question, matching the format 'Answer: Yes' or 'Answer: No'. Then, explain your reasoning. Does the pathology report indicate that the patient has any type of adenoma, adenomatous/dysplastic lesion(s), or dysplasia of any grade in any colon or rectal sample? Exclude sessile serrated adenomas unless they are specified to have dysplasia.*
*<<<*
*Pathology report:*
*{Insert Microscopic Exam text or full-text pathology report}*
*>>>*
*Does the pathology report indicate that the patient has any type of adenoma (excluding sessile serrated adenoma), adenomatous/dysplastic lesion(s), or dysplasia of any grade in the colon or rectum?*
*Answer:”*

***Indefinite for dysplasia ascertainment***
We kept the initial search terms the same as the dysplasia terms for targeting “indefinite for dysplasia”. As stated above, these are inclusive of all the below terms:

Invasive CRC search terms: ‘%carcinoma%’, ‘%tumor%’, ‘%invasi%’
HGD/CRC search terms: ‘%HGD%’, ‘%high-grade%’, ‘%high grade%’, ‘%in situ%’, ‘%in-situ%’, ‘%intramucosal%’
Additional dysplasia search terms: ‘%dysplas%’, ‘%lgd%’, ‘%low grade%’, ‘%low-grade%’, ‘%dalm%’, ‘%adenoma%’

Keeping the same format, the prompt for identifying “indefinite for dysplasia” is as follows:

"*The text provided is a pathology report. Answer yes or no to the following question, matching the format 'Answer: Yes' or 'Answer: No'. Then, explain your reasoning. Does the pathology report indicate that any colon or rectal sample contains tissue which is considered ‘indefinite for dysplasia’? ‘Indefinite for dysplasia’ is defined as uncertainty about the presence of dysplasia and may include descriptions of equivocal findings which are insufficient to confirm or exclude dysplasia. Answer yes if any colon or rectal sample is considered indefinite for dysplasia.*

*<<<*
*Pathology report:*
*{Insert Microscopic Exam text or full-text pathology report}*

>>>

*Does the pathology report indicate that the patient has any colon or rectal sample that is indefinite for dysplasia?*

*Answer:"*

There are several details to note, which are relevant for all of our LLM use-cases:

1. We force the model's output to be " Yes" or " No" using a "grammar", which is a feature of llama.cpp[11]. Any tokens not matching this rule will not be considered. Generation is terminated immediately after "Yes" or "No" is produced, reducing unnecessary and expensive generation. We include our simple grammar in our HuggingFace repository.
2. We initiate the model's response with "Answer:" as a part of the prompt, which effectively forces the model's response to begin with "Answer:". LLMs often begin with phrases like "In this pathology report…" or "In this case…", which makes it unnatural for the response to match the grammar ("Yes" or "No"). When we begin with "Answer:" the next token is naturally "Yes" or "No" based on the instructions in the prompt. We also found better performance when starting with "Answer:" as opposed to including a trailing space "Answer:[space]" because tokens are typically "[space][word]" as opposed to "[word][space]".
3. We ask the model to "explain your reasoning" but do not actually give it the opportunity to explain. In testing, this seemed to perform better than not asking it to explain, but that may not always be the case. Modifying the grammar to allow the model to explain could help to identify the error of the model's reasoning in development.
4. Exact formatting of the prompt, including preceding headers (such as "<|start_header_id|>system<|end_header_id|>" for Llama-3) were included based on the guidelines given by the instruct-tuned model information provided by Meta, Mistral, and Google, respectively. For all models, the text that remained constant before the Pathology report text (up to and including "<<<\nPathology report: " was cached for efficiency, and is denoted as the "system prompt" in the code.
5. Temperature is always set to 0 for reproducibility, which forces the model to always select the highest probability token that satisfies the grammar. Probabilities of the model returning "Yes" and "No" can also be extracted if desired. This may be helpful for identifying more difficult cases to be manually annotated or fed to a larger model.

Note also that it may be more efficient to run a single query with all dysplasia search terms through the LLM, and then ask if there is invasive cancer, high grade dysplasia or adenocarcinoma, any dysplasia, or none. However, we did not implement this and there may be costs to accuracy in switching from a binary "Yes" or "No" to four possible outputs. As with LLMs in general, there will be tradeoffs between computational cost and accuracy. In our case, the relatively small number of pathology reports fin the CRC and HGD/CRC plausible sets made it feasible to run these tasks separately, even while restricted to CPU-only inference.

***Model validation details***

We performed validation only in the set of notes that passed our regular expression filters. Considering the very low expected prevalence (potentially zero) outside of the regular expression filters, this approach provides a more informative assessment of the LLMs' performance, as we are considerably more likely to include some false negative cases in our validation.

Validation was performed at the level of the pathology report, consistent with the LLM prompt asking if the given features are present in any colon or rectal sample. We were evaluating presence or absence of the given feature, not the most advanced lesion. As such, dysplasia and CRC could both be validated as “Yes” for a given note if both features were present. Similarly, indefinite for dysplasia and dysplasia could also both be ‘Yes’ in the same report. Validation was performed independently for each of the four tasks, even if notes overlapped in development or validation sets across tasks.

Either N=100 (CDW) or N=150 (MVP) randomly selected putative positive cases and the same number of putative negative cases were selected for review. The randomly selected putative positive and negatives were combined and shuffled, then written to a csv and validated in Excel. Putative positive (negative) cases were defined as cases where Llama-3 responded “Yes” (“No”). An exception is IBD-CRC in MVP, where the total number of positives by either model was 159 in the entire cohort, so we validated all 159 which were positive by either model. This led to 307 total validated reports, as two of the nine additional reports deemed positive by either model were included in the original development set of 150 putative negatives, as they were negative by Llama-3 and positive by Gemma-2.

Metastatic adenocarcinoma suspected or known to be from sites other than the colon or rectum were considered negative even if found in the colon or rectum. As discussed, sessile serrated adenoma/lesion/polyp were considered negative for the dysplasia algorithm unless explicitly stated to include dysplasia. Indefinite for dysplasia was validated as “No” for the dysplasia algorithm in IBD, consistent with our validation approach which required definitive diagnosis. Similarly, “cannot rule out invasive adenocarcinoma”, “bordering on high-grade dysplasia” and other statements conveying uncertainty were validated as “No” for their respective diagnoses. For the CRC algorithm, invasive adenocarcinoma was required for “Yes” validation, which was defined as T stage greater than or equal to 1, or equivalent language. “Adenocarcinoma” without further qualifying language was assumed to imply invasion. For the dysplasia algorithm, we note that only low-grade and high-grade dysplasia were counted as positive (see Main text Figure 1). Carcinoma in-situ, intramucosal adenocarcinoma, and invasive adenocarcinoma were not validated as “Yes” for the dysplasia algorithm unless they were specified to arise in a dysplastic lesion or there were additional findings which included dysplasia. While this distinguishes very similar concepts, HGD and carcinoma in-situ, this is mostly an irrelevant issue as the HGD/CRC algorithm identifies both as positive regardless. In IBD, indefinite for dysplasia was only validated as “Yes” when the pathologist expressed uncertainty about the diagnosis of dysplasia, which often included the terms ‘indefinite for dysplasia’ or ‘indeterminate for dysplasia’. Confirmed dysplasia would be validated as “No” for the indefinite for dysplasia task.

The Microscopic Exam section was used to validate the LLM responses. However, when validating the full pathology reports, the full pathology report was used in cases where the models disagreed. The full pathology report validation differed from the Microscopic Exam validation 0 of the 15 times when either of the models disagreed for CRC (N=227 total notes), 2 of 26 times when models disagreed for HGD/CRC (N=235 total reports), and 0 of 4 times when models disagreed for Dysplasia (N=239 total notes). See Supplementary Table S6 for details.

#### *Calculating performance metrics*

We calculated performance metrics as follows, where $w$ = prevalence of model positives in the plausible set:

$$Sensitivity = PPV * w/(PPV * w + (1 - NPV) * (1 - w))$$
$$Specificity = NPV * (1 - w)/(NPV * (1 - w) + (1 - PPV) * w)$$
$$F1 = 2 * PPV * Sensitivity/(PPV + Sensitivity)$$
$$MCC = \sqrt{PPV * Sensitivity * Specificity * NPV} - \sqrt{(1 - PPV) * (1 - Sensitivity) * (1 - Specificity) * (1 - NPV)}$$

Because we use Llama-3 for conditioning/stratifying our validation, the calculation of the performance metrics using other models, such as Gemma-2, must explicitly incorporate this conditioning. For example, the definition of the positive predictive value (PPV) is P(R|M) where we define R as review positive and M as model positive. In the case where Llama-3 is the model, and Llama-3 was used to condition, PPV is a straightforward calculation of the number of TP in the reviewed set divided by the number of positives reviewed (TP + FP). However, our conditioning on Llama-3 means that we do not have an estimate of P(R|M) directly when the model is Gemma-2. Instead, we know P(R|GL) and P(R|GL') which are, respectively, the probability of review positive given that gemma and llama are both positive, and the probability of review positive given that gemma is positive and llama is negative. Therefore, our conditioning requires adjustments to estimate the performance metrics of PPV, NPV, which affect the downstream estimates of sensitivity, specificity, F1-score, and MCC. Building off of Appendix A of Liu et al.[14], we show the estimates below, for a given model M, review R, and conditional/stratifying model L (for Llama-3). M, R, L, denote positive and M', R', and L' denote negative. These capture all possibilities because the variables are binary. Equations that we can get directly from our data will be bolded.

First, expanding by the law of total probability, we have

$$PPV\ (precision) = P(R|M) = \boldsymbol{P(R|M,L)P(L|M)} + \boldsymbol{P(R|M,L')P(L'|M)}$$

If we can estimate $P(L|M)$, we can compute this directly from our data. For example, we have run both Llama-3 and Gemma-2 on N path reports without stratification, and we can see how often Llama-3 is positive when Gemma-2 is positive. However, for some models we do not have this information, requiring us to estimate PPV in a different way. Next, applying Bayes' rule to $P(R|M,L)$ and $P(R|M,L')$

$$PPV = \frac{P(R, M|L)}{P(M|L)} P(L|M) + \frac{P(R, M|L')}{P(M|L')} P(L'|M)$$

$$PPV = \frac{P(R, M|L)P(L|M)}{P(M|L)} + \frac{P(R, M|L')P(L'|M)}{P(M|L')}$$

Again, applying Bayes' rule to $P(L|M)$, $P(M|L)$, $P(L'|M)$, and $P(M|L')$:

$$PPV = P(R, M|L) * \frac{P(L, M)}{P(M)} * \frac{P(L)}{P(M, L)} + P(R, M|L') * \frac{P(L', M)}{P(M)} * \frac{P(L')}{P(M, L')}$$

Simplifying:

$$PPV = \frac{P(R, M|L)P(L) + \ P(R, M|L')P(L')}{P(M)}$$

Finally, replacing $P(M)$ with $P(M|L)P(L) + P(M|L')P(L')$ using the law of total probability, which gives us a quantity we can directly calculate from our data:

$$\boldsymbol{w = P(M) = P(M|L)P(L) + P(M|L')P(L')}$$

$$\boldsymbol{Precision \ = \ PPV = \frac{P(R, M|L)P(L) + \ P(R, M|L')P(L')}{w}}$$

We can calculate $w$ either directly based on the total number of model positives in the plausible set of pathology reports or based on the equation above when we have only run the model on the validation set. In general, we then use $w$ in place of $P(M)$.

Next, we calculate the negative predictive value (NPV). Starting with a similar expansion using the law of total probability:

$$NPV \ = \ P(R'|M') \ = \ \boldsymbol{P(R'|M', L)P(L|M') + P(R'|M', L')P(L'|M')}$$

Again, this can be computed directly from our data in the cases where we know the prevalence of L and M. In other cases, we continue, applying Bayes rule to $P(R'|M', L)$ and $P(R'|M', L')P$:

$$NPV = \frac{P(R', M'|L)}{P(M'|L)} P(L|M') + \frac{P(R', M'|L')}{P(M'|L')} P(L'|M')$$

Now applying Bayes rule to $P(L|M')$, $P(M'|L)$, $P(L'|M')$, and $P(M'|L')$:

$$NPV = P(R', M'|L) * \frac{P(L, M')}{P(M')} * \frac{P(L)}{P(M', L)} + P(R', M'|L') * \frac{P(L', M')}{P(M')} * \frac{P(L')}{P(M', L')}$$

Simplifying:

$$NPV = \frac{P(R', M'|L)P(L) + P(R', M'|L')P(L')}{P(M')}$$

$$\boldsymbol{NPV} = \frac{\boldsymbol{P(R', M'|L)P(L) + P(R', M'|L')P(L')}}{\boldsymbol{(1 - w)}}$$

Which gives us a quantity we can estimate directly from our data even if we do not know the prevalence of the model in the larger set of path reports, as long as we can calculate $w$.

Now, restating the derivation from Liu et al.[14], where $w$ is the prevalence of the model M:

$$sensitivity\ (recall) = P(M|R) = \frac{P(M, R)}{P(R)} = \frac{P(R|M)P(M)}{P(R)}$$

$$sensitivity\ (recall) = \frac{PPV * w}{P(R|M)P(M) + P(R|M')P(M')}$$

$$sensitivity\ (recall) = \frac{PPV * P(M)}{PPV * P(M) + (1 - NPV) * P(M')}$$

$$sensitivity\ (recall) = \frac{\boldsymbol{PPV * w}}{\boldsymbol{PPV * w + (1 - NPV) * (1 - w)}}$$

And restating the derivation for specificity from Liu et al.[14]:

$$specificity = P(M'|R') = \frac{P(R', M')}{P(R')}$$

$$specificity = \frac{P(R'|M')P(M')}{P(R')}$$

$$specificity = \frac{NPV * (1 - w)}{P(R'|M)P(M) + P(R'|M')P(M')}$$

$$specificity = \frac{\boldsymbol{NPV * (1 - w)}}{\boldsymbol{(1 - PPV) * w + NPV * (1 - w)}}$$

Finally, the downstream metrics, F1 and MCC, can be calculated given the estimated PPV, NPV, specificity, and sensitivity.

$$\boldsymbol{F1} = \frac{\boldsymbol{2 * PPV * sensitivity}}{\boldsymbol{PPV + sensitivity}}$$

$$\boldsymbol{MCC} = \sqrt{\boldsymbol{PPV} * \boldsymbol{sensitivity} * \boldsymbol{specificity} * \boldsymbol{NPV}} - \sqrt{(\mathbf{1} - \boldsymbol{PPV}) * (\mathbf{1} - \boldsymbol{sensitivity}) * (\mathbf{1} - \boldsymbol{specificity}) * (\mathbf{1} - \boldsymbol{NPV})}$$

It is known that the some of these metrics are directly dependent on the prevalence of model positives, also known as the class ratio[15]. Because we see such large differences in the class imbalance, especially for indefinite for dysplasia, we also compute the calibrated precision ($Prec_c$, equivalent to calibrated PPV) and calibrated F1-score, $F1_c$[15]. For these, we arbitrarily set the reference class ratio $\pi_0 = 0.5$ and use $\pi = w$ for the model prevalence.

$$PPV_c = Prec_c = \frac{TP}{TP + \frac{\pi(1-\pi_0)}{\pi_0(1-\pi)}} = \frac{1}{1 + \frac{\pi(1-\pi_0)FP}{\pi_0(1-\pi)TP}}$$

Substituting $w = \pi$, we rewrite as:

$$PPV_c = Prec_c = \frac{1}{1 + \frac{w(1-\pi_0)FP}{\pi_0(1-w)TP}}$$

We estimate the number of false positives in a given dataset as:

$$\boldsymbol{FP} = (\mathbf{1} - \boldsymbol{PPV})\boldsymbol{w}$$

And the number of TP as:

$$\boldsymbol{TP} = \boldsymbol{PPV} * \boldsymbol{w} + (\mathbf{1} - \boldsymbol{NPV})(\mathbf{1} - \boldsymbol{w})$$

Giving us a calibrated precision of:

$$\boldsymbol{PPV_c} = \boldsymbol{Prec_c} = \frac{\mathbf{1}}{\mathbf{1} + \frac{\boldsymbol{w}^{\mathbf{2}}(\mathbf{1} - \boldsymbol{\pi_0})(\mathbf{1} - \boldsymbol{PPV})}{\boldsymbol{\pi_0}(\mathbf{1} - \boldsymbol{w})[\boldsymbol{PPV} * \boldsymbol{w} + (\mathbf{1} - \boldsymbol{NPV})(\mathbf{1} - \boldsymbol{w})]}}$$

The calibrated F1-score is simply calculated with calibrated precision used in place of precision (PPV):

$$\boldsymbol{F1_c} = \frac{\mathbf{2} * \boldsymbol{Prec_c} * \boldsymbol{sensitivity}}{\boldsymbol{Prec_c} + \boldsymbol{sensitivity}}$$

We used bootstrapping to determine the confidence intervals. Specifically, for each subset we uniformly re-sampled the validated dataset with replacement, keeping the size of the dataset the same. Then, we calculated the given metric, accounting for conditional probabilities as noted above. We repeated this process 10,000 times to build a distribution for the metric. To construct a 95% confidence interval, we take the 2.5th percentile and the 97.5th percentile of this distribution, using the *quantile()* function in R. Since bootstrapping is only informative when there is some variability in the data (e.g., when PPV is not exactly 1), we also computed the analytical binomial confidence interval for NPV and PPV. We used the direct binomial confidence interval for Llama-3 results but for Gemma-2, due to conditioning, we approximated the binomial confidence interval by setting the number of successes $k = round(PPV * n)$, where $n$ is the number of model positives that were validated for a given model and task. Then we used the *binom.test* R function to determine 95% confidence intervals. For each bound (lower and upper), we took the more conservative of the two intervals, between bootstrapping and analytical, and report that as the bound for PPV and NPV.

***Lessons learned from applying LLMs in structured and unstructured pathology report data***

The 48 reports chosen for prompt development had Microscopic Exam sections that contained the term ‘carcinoma’ and were drawn randomly without consideration for the IBD diagnosis status of the patient. From manual chart review, 16 had invasive colorectal adenocarcinoma, 16 had high-grade dysplasia or adenocarcinoma in the colon or rectum, and 16 had neither. Some of the 48 also had dysplasia that was not high-grade in the colon or rectum. Prompt iteration and model selection occurred mainly in these 48, which were excluded from all future validation sets. Minor changes were made iteratively to the prompts to correct any obvious errors related to the prompt (e.g., missing tubular adenomas before dysplasia prompt included 'any adenoma') using additional development sets for each task, which consisted of 818 reports for CRC, 200 reports for HGD/CRC, and 572 reports for any dysplasia.

For the diagnosis task for ‘indefinite for dysplasia’ in IBD, we skipped the development set iteration step to demonstrate the adaptability of our approach. Instead of intensive manual review, we based the prompt on discussions with expert gastroenterologists and pathologist (SCS, SG, LJJ). We agreed that the prompt was fixed before running the LLMs and then validated the results in the same way as for the other tasks. We were confident in doing this because the general format of the prompt could be kept the same, with modifications as necessary to fit the new concept.

*Dysplasia prompt iteration*

When working with smaller models (< 30B parameters), it is important to be specific and direct in the prompt. For example, the original question for the dysplasia prompt simply asked:

*"Does the pathology report indicate that the patient has adenocarcinoma, adenomatous lesions, or dysplasia of any grade in any colon or rectal sample?"* (v1)

When the model missed many instances of the most common dysplastic lesion, tubular adenomas, we iterated to explicitly include “any adenoma”. This took a few different forms as we iterated on the development set to finalize the prompt.

*"Does the pathology report indicate that the patient has adenocarcinoma, adenomatous lesions (including any adenomas), or dysplasia of any grade in any colon or rectal sample?"* (v2)

*“Does the pathology report indicate that the patient has adenocarcinoma, adenomatous lesions, any adenoma, or dysplasia of any grade in any colon or rectal sample?”* (v3)

*“Does the pathology report indicate that the patient has adenocarcinoma, adenomatous lesion(s), any type of adenoma, or dysplasia of any grade in any colon or rectal sample?”* (v4)

Similarly, to reduce the possibility of the model picking up on adenocarcinoma from a non-colorectal specimen, and because we have a separate prompt for identifying CRCs, we removed the term “adenocarcinoma”. As a result, the dysplasia prompt did not include dysplasia *and any more advanced lesion*. The overlap of the targets of each prompt is shown in main figure 1. This change led to the prompt:

*“Does the pathology report indicate that the patient has any type of adenoma, adenomatous/dysplastic lesion(s), or dysplasia of any grade in any colon or rectal sample?”* (v5)

Another issue arises in this prompt due to unclear historical terminology used in pathology reports. ‘Sessile Serrated Adenomas’, now referred to more commonly as ‘Sessile Serrated Lesions’, unlike other adenomas, do not inherently contain dysplasia unless explicitly stated. Therefore, in order to identify dysplasia, the phrase “any type of adenoma” must be conditioned, leading to the final question:

*"Does the pathology report indicate that the patient has any type of adenoma, adenomatous/dysplastic lesion(s), or dysplasia of any grade in any colon or rectal sample? Exclude sessile serrated adenomas unless they are specified to have dysplasia."* (v6 and FINAL)

The question above is the final one incorporated into the larger prompt shown in supplementary section *“Dysplasia Ascertainment”*. While not as extensive as the effort required to develop rules-based NLP approaches, prompt iterating does require some human effort. There was a similar iteration process for identifying adenocarcinoma and ensuring that the report identified invasive (T stage greater than or equal to 1) adenocarcinoma. The prompting for HGD/CRC was more straightforward, as shown below.

*HGD/CRC prompt iteration*

Following some prompt iteration using the original 48 pathology reports, there was only one change made after development set validation for the HGD/CRC prompt.

*“Does the pathology report indicate that the patient has adenocarcinoma (including in-situ or intramucosal adenocarcinoma) or high-grade dysplasia in any colon or rectal sample?"* (v1)

This was simplified to the following:

*“Does the pathology report indicate that the patient has adenocarcinoma or high-grade dysplasia in any colon or rectal sample?"* (v2 and FINAL)

*CRC prompt iteration*

Following prompt iteration using the original 48 pathology reports, there was only one change made after development set validation for the CRC prompt.

*"For the purpose of this query, colorectal cancer is defined as the presence of malignant carcinoma in any colon or rectal sample. Conditions such as high-grade dysplasia, in-situ carcinoma, or intramucosal carcinoma in the absence of malignant carcinoma are not classified as colorectal cancer in this context. Based on these criteria, does the note indicate that the patient has colorectal cancer? Answer Yes or No."*

We refined this prompt to use more specific terms that were more relevant in the context of pathology reports. We replaced "malignant" with "invasive", and made sure to specify that "adenocarcinoma" was the target, instead of keeping the more general terms "carcinoma" and "colorectal cancer". This was changed to the following:

*"We are interested in identifying whether invasive adenocarcinoma (stage greater than or equal to 1) is present in *any* colon or rectal sample. Without definite invasion identified, conditions such as 'high-grade dysplasia', 'in-situ [adeno]carcinoma', or 'intramucosal [adeno]carcinoma' are not typically classified as invasive adenocarcinoma. If the sample is classified as having adenocarcinoma without further specification, this typically implies invasive adenocarcinoma. Answer yes or no to the following question, matching the format 'Answer: Yes' or 'Answer: No'. Then, explain your reasoning. Does the pathology report indicate that the patient has an invasive adenocarcinoma in any colon or rectal sample?"* (v2 and FINAL)

As with any task using LLMs, there will be a tradeoff between the size (quality, cost) of the model, and the need to perform prompt iteration and preprocessing. Using state-of-the-art larger models such as Llama3-405B[5], GPT-4o or others, we could probably have kept the initial dysplasia prompt and let the model determine that tubular adenomas inherently contain dysplasia whereas sessile serrated adenomas do not. Further, we would expect that the larger models handle full-text pathology reports better than smaller ones, possibly reducing the need for the Pathology Domain sectioning. Even since our testing, an updated version of Llama-3, "Llama-3.1-8B-Instruct", has been released with additional longer context training. This newer model may address some of the shortcomings we found in applying Llama-3 to full pathology reports. However, computational cost can still be saved by these preprocessing steps, especially those like sectioning that reduce the input text required.

There was not a clear performance advantage in general to using two models compared to the better performing model, which in our case was Gemma-2-SPPO (see Supplementary Table S5). One obvious exception is that using two models will allow those trying to adapt their approach to target either higher sensitivity (either model says "Yes") or specificity (both models say "Yes"). It appears that, if the resources allow, simply using a better model would be advantageous to using two smaller models. However, it may be useful to use two models for prompt iteration, as it is good to identify problems with the prompt (where both models get it wrong) vs. problems with the model (only one model gets it wrong). As in our case (2 compute resource-constrained workspaces), it is often easier and faster to run two models concurrently instead of one larger model.

#### *Expanding the regular expression set for dysplasia*

Possible typos added to ‘adenoma’ and ‘carcinoma’, in addition to other terms associated with dysplasia, added to augment the regular expression (regex) query did not return many additional notes.

Original regex set: ‘%carcinoma%’, ‘%tumor%’, ‘%invasi%’, ‘%HGD%’, ‘%high-grade%’, ‘%high grade%’, ‘%in situ%’, ‘%in-situ%’, ‘%intramucosal%’, ‘%dyspas%’, ‘%lgd%’, ‘%low grade%’, ‘%low-grade%’, ‘%dalm%’, ‘%adenoma%’

Expanded regex set: '%dysplas%', '%lgd%', '%low grade%', '%low-grade%', '%low_grade%', '%dalm%', '%adenoma%', '%adneoma%', '%ademona%', '%adnoma%', '%adeoma%', '%adenmoa%', '%adenoms%', '%adenona%', '%adenome%', '%ademoma%', '%adnema%', '%invasi%', '%tumor%', '%tumour%', '%carcinoma%', '%carcioma%', '%carcinomia%', '%carcimona%', '%carincoma%', '%carcnoma%', '%carcimona%', '%carcinom%', '%carcinmoa%', '%carinoma%', '%high grade%', '%hgd%', '%high-grade%' , '%high_grade%', '%in-situ%', '%in situ%', '%intramucosal%', '%hyperchromasia%', '%pseudostratification%' , '%crowding%', '%nuclear enlargement%', '%elongation%', '%increased mitot%'

There were originally 14,379 unique dysplasia notes for IBD-Dysplasia in MVP. The expanded search terms return 14,393 unique notes. All 14 were checked and none of the 14 contain a diagnosis of dysplasia. While this still does not rule out false negatives occurring outside the regex set, it increases confidence that the regular expression search is performing well.

#### *CRC metrics at low prevalence (without filtering)*

Class imbalance represents a challenge in identifying rare diseases from notes and evaluating performance. Our regular expression (regex) filtering removed many assumed negative reports without feeding them to the LLM. This represents a removal of a large number of notes which may, if fed to the LLM, result in false positives. We applied our IBD-CRC algorithm in both LLMs to N=21,372 Microscopic Exam sections in MVP that did not undergo any CRC-specific regular expression filtering. That is, these pathology reports were identified as “Colonoscopy path reports” (see Figure 1) by filtering on the Specimen section but were not filtered further. In the 160 reports where either LLM answered “Yes”, all but one were within the CRC regular expression set, matching to ‘%tumor%’, ‘%carcinoma%’, or ‘%invasive%’. This one report that did not match was a false positive and was only considered a “Yes” by Gemma-2, not Llama-3. Thus, only one false positive occurred outside the regular expression set, and the positive predictive value we report within the regex set remains approximately accurate even in the imbalanced case (<1% prevalence). This suggests that the regular expression filtering in CRC is working as intended, reducing computational expense without compromising accuracy. We cannot rule out the possibility of false negatives outside the regex set, but we would likely need to validate far too many reports to find one. In the case of dysplasia, we expanded the regex set to include typos and other phrases for the dysplasia regular expressions and found that all added notes were true negatives (see Supplementary section “*Expanding the regular expression set for dysplasia”*).

*Cloning llama.cpp and running inference*

The following steps can be used to clone llama.cpp and run inference in relatively few steps. These instructions are also available at https://github.com/ggml-org/llama.cpp

```
git clone https://github.com/ggml-org/llama.cpp
cd llama.cpp
cmake -B build
cmake –build build –config Release
./build/bin/llama-cli -m your_model.gguf -p "I believe the meaning of life is" -n 128
```

While this represents a first step to perform simple inference, a more tailored approach is desired for the purposes of structuring data. Specifically, caching the system prompt while running many inputs (e.g. pathology reports) through the same task. We modify the example given in llama.cpp/examples/parallel, which does this caching inherently while allowing users to input their own prompt file. Our modification changes the system prompt and input prompts based on the task we specify, which is added as a parameter "--extractionType crc". Our code is available at https://github.com/bdj34/llama.cpp_data_extraction

With the rapid development of llama.cpp, we have continued to merge new updates to our repo. However, it may be advisable to modify your own version of https://github.com/ggml-org/llama.cpp rather than use ours, as merge conflicts do occur and must be resolved manually.

### Supplementary Tables

| Overview | Source | Cohort | Report filters | N distinct path reports | N distinct patients |
|---|---|---|---|---|---|
| All MVP patients | MVP | All | N/A | N/A | 913318 |
| MVP patients in IBD cohort | MVP | IBD | N/A | N/A | 12682 |
| MVP patients in non-IBD cphort | MVP | Non-IBD | N/A | N/A | 887478 |
| All MVP pathology reports | MVP Path Domain | All | All path reports | 2600043 | 604311 |
| All Partitioned (matching colon specimen terms) | MVP Path Domain | All | All colonoscopy path reports | 279964 | 170806 |
| All IBD (matching colon specimen terms) | MVP Path Domain | IBD | All colonoscopy path reports | 21372 | 6321 |
| All Non-IBD (matching colon specimen terms) | MVP Path Domain | Non-IBD | All colonoscopy path reports | 249823 | 159675 |
| IBD-CRC | MVP Path Domain | IBD | CRC | 607 | 463 |
| IBD-HGD/CRC | MVP Path Domain | IBD | HGD/CRC | 1492 | 1024 |
| IBD-Dysplasia | MVP Path Domain | IBD | Dysplasia | 14379 | 5101 |
| Non-IBD-CRC | MVP Path Domain | Non-IBD | CRC | 12806 | 9290 |
| Non-IBD-HGD/CRC | MVP Path Domain | Non-IBD | HGD/CRC | 36286 | 26524 |
| Non-IBD-Dysplasia | MVP Path Domain | Non-IBD | Dysplasia | 184711 | 121917 |
| All CDW patients | CDW | All | N/A | N/A | 15216068 |
| CDW patients in IBD cohort | CDW | IBD | N/A | N/A | 102447 |
| CDW patients in non-IBD cphort | CDW | Non-IBD | N/A | N/A | 15037696 |
| All CDW pathology reports | CDW Path Domain | All | All path reports | 16335204 | 4972551 |
| All Colonoscopy-related (matching colon specimen terms) | CDW Path Domain | All | All colonoscopy path reports | 2899321 | 1834930 |
| All IBD (matching colon specimen terms) | CDW Path Domain | IBD | All colonoscopy path reports | 182500 | 60011 |
| All Non-IBD (matching colon specimen terms) | CDW Path Domain | Non-IBD | All colonoscopy path reports | 2649236 | 1737946 |
| IBD-CRC | CDW Path Domain | IBD | CRC | 3825 | 2904 |
| IBD-HGD/CRC | CDW Path Domain | IBD | HGD/CRC | 7721 | 5535 |
| IBD-Dysplasia | CDW Path Domain | IBD | Dysplasia | 65622 | 28179 |
| Non-IBD-CRC | CDW Path Domain | Non-IBD | CRC | 104708 | 78681 |
| Non-IBD-HGD/CRC | CDW Path Domain | Non-IBD | HGD/CRC | 224972 | 172255 |
| Non-IBD-Dysplasia | CDW Path Domain | Non-IBD | Dysplasia | 1023245 | 721656 |

**Supplementary Table S1: Cohorts and search term filtering of pathology reports.** Numbers of distinct pathology reports and patients for each of the filtering steps in MVP (top, white) and CDW (bottom, light gray). Path Domain = Pathology Domain.

| Method | Cohort | Data Source | Task | Compute | Model | Input type | Number of reports | Mean number of characters per input | Total time (seconds) | Time per note (seconds) | Time per input char (NOT TOKEN) |
|---|---|---|---|---|---|---|---|---|---|---|---|
| This work (LLMs) | IBD-colitis | MVP | Dysplasia | CPU | llama-3-8B | Microscopic exam | 300 | 912.6 | 13662.1 | 45.5 | 0.050 |
| This work (LLMs) | IBD-colitis | MVP | Dysplasia | CPU | gemma-2-9B | Microscopic exam | 300 | 912.6 | 16138.8 | 53.8 | 0.059 |
| This work (LLMs) | No IBD-colitis | MVP | Dysplasia | CPU | llama-3-8B | Microscopic exam | 1636 | 587.8 | 47093 | 28.8 | 0.049 |
| This work (LLMs) | No IBD-colitis | MVP | Dysplasia | CPU | gemma-2-9B | Microscopic exam | 300 | 587.8 | 12251.3 | 40.8 | 0.069 |
| This work (LLMs) | IBD-colitis | MVP | Dysplasia | CPU | llama-3-8B | Full pathology report | 253 | 4849.6 | 59059.1 | 233.4 | 0.048 |
| This work (LLMs) | IBD-colitis | MVP | Dysplasia | CPU | gemma-2-9B | Full pathology report | 253 | 4849.6 | 78011.3 | 308.3 | 0.064 |
| This work (LLMs) | IBD-colitis | MVP | Dysplasia | CPU | llama-3.2-3B | Microscopic exam | 300 | 912.6 | 5149.9 | 17.2 | 0.019 |
| This work (LLMs) | IBD-colitis | MVP | Dysplasia | CPU | gemma-2-2B | Microscopic exam | 300 | 912.6 | 3831.7 | 12.8 | 0.014 |
| This work (LLMs) | IBD-colitis | MVP | IND | GPU | llama-3-8B | Microscopic exam | 14379 | 999.2 | 1551.5 | 0.11 | 0.00011 |
| This work (LLMs) | IBD-colitis | MVP | IND | GPU | gemma-2-9B | Microscopic exam | 14379 | 999.2 | 2057.6 | 0.14 | 0.00014 |
| **Published rule-based approaches** | | | | | | | | | | | |
| Benson et al. (2023) "Leveraging…" | General population | | All features | | | Full pathology report | 7200 | | 60 | 0.008 | |

**Supplementary Table S2: Run times for all models, comparing CPU, GPU, and published approaches.** Full note is significantly slower than using Microscopic Exam only, as expected. As run time is mostly a function of input text length and model size, Gemma-2-9B is slightly slower than Llama-3-8B for the same input, as expected (~9B params vs. ~8B params). Models process text in units of tokens, not characters, but tokenization differs across models. For that reason, we show the time per input character, which will correlate well with the number of tokens. nchar() function in R used to calculate the number of characters in the input text. Time per note colored using standard green-yellow-red gradient scale, from lowest value (green) to highest value (red), with yellow at midpoint. Time per input character in white-red scale from lowest value (white) to highest value (red). IBD = Inflammatory Bowel Disease; IND = Indefinite for dysplasia; CPU = 4-core, 2.3 GHz Intel Haswell, no TSX; GPU = 1 Nvidia A40.

| Task | Cohort | Source | Input | Model prevalence estimate | PPV (LB - UB) | NPV (LB - UB) | PPVc | Recall (Sensitivity) | Specificity | F1 | F1c | MCC | Cohen's kappa |
|---|---|---|---|---|---|---|---|---|---|---|---|---|---|
| CRC | IBD | MVP | Microscopic exam | 0.250 | 0.947 (0.90 - 0.98) | 0.961 (0.92 - 0.99) | 0.984 | 0.891 | 0.982 | 0.918 | 0.935 | 0.891 | 0.889 |
| CRC | IBD | CDW | Microscopic exam | 0.262 | 0.960 (0.90 - 0.99) | 0.980 (0.93 - 1.00) | 0.986 | 0.945 | 0.986 | 0.952 | 0.965 | 0.935 | 0.910 |
| CRC | IBD | MVP | Full pathology report | 0.251 | 0.895 (0.82 - 0.96) | 0.941 (0.88 - 0.98) | 0.968 | 0.835 | 0.964 | 0.864 | 0.897 | 0.817 | 0.877 |
| CRC | non-IBD | MVP | Microscopic exam | 0.515 | 0.980 (0.94 - 1.00) | 0.887 (0.82 - 0.94) | 0.981 | 0.902 | 0.977 | 0.939 | 0.940 | 0.873 | 0.920 |
| CRC | non-IBD | CDW | Microscopic exam | 0.535 | 0.970 (0.91 - 1.00) | 0.940 (0.87 - 0.98) | 0.967 | 0.949 | 0.965 | 0.959 | 0.958 | 0.912 | 0.940 |
| HGD/CRC | IBD | MVP | Microscopic exam | 0.149 | 0.967 (0.92 - 0.99) | 0.967 (0.92 - 0.99) | 0.995 | 0.836 | 0.994 | 0.897 | 0.909 | 0.880 | 0.947 |
| HGD/CRC | IBD | CDW | Microscopic exam | 0.186 | 0.950 (0.89 - 0.99) | 0.960 (0.90 - 0.99) | 0.990 | 0.844 | 0.988 | 0.894 | 0.911 | 0.870 | 0.970 |
| HGD/CRC | IBD | MVP | Full pathology report | 0.141 | 0.923 (0.82 - 0.99) | 0.961 (0.92 - 0.99) | 0.989 | 0.795 | 0.987 | 0.854 | 0.882 | 0.832 | 0.941 |
| HGD/CRC | non-IBD | MVP | Microscopic exam | 0.260 | 0.993 (0.96 - 1.00) | 0.927 (0.87 - 0.97) | 0.998 | 0.826 | 0.997 | 0.902 | 0.904 | 0.871 | 0.947 |
| HGD/CRC | non-IBD | CDW | Microscopic exam | 0.370 | 1.000 (0.96 - 1.00) | 0.970 (0.91 - 1.00) | 1.000 | 0.951 | 1.000 | 0.975 | 0.975 | 0.961 | 0.950 |
| Dysplasia | IBD | MVP | Microscopic exam | 0.214 | 0.973 (0.93 - 0.99) | 0.987 (0.95 - 1.00) | 0.993 | 0.952 | 0.993 | 0.963 | 0.972 | 0.952 | 0.920 |
| Dysplasia | IBD | CDW | Microscopic exam | 0.310 | 0.960 (0.90 - 0.99) | 0.990 (0.95 - 1.00) | 0.982 | 0.977 | 0.982 | 0.969 | 0.980 | 0.955 | 0.950 |
| Dysplasia | IBD | MVP | Full pathology report | 0.208 | 0.991 (0.95 - 1.00) | 0.987 (0.94 - 1.00) | 0.998 | 0.953 | 0.998 | 0.972 | 0.975 | 0.965 | 0.967 |
| Dysplasia | non-IBD | MVP | Microscopic exam | 0.895 | 0.987 (0.95 - 1.00) | 0.973 (0.93 - 0.99) | 0.897 | 0.997 | 0.895 | 0.992 | 0.944 | 0.926 | 0.953 |
| Dysplasia | non-IBD | CDW | Microscopic exam | 0.875 | 0.990 (0.95 - 1.00) | 0.980 (0.93 - 1.00) | 0.934 | 0.997 | 0.933 | 0.994 | 0.965 | 0.950 | 0.920 |
| IND | IBD | MVP | Microscopic exam | 0.023 | 0.800 (0.73 - 0.86) | 0.993 (0.96 - 1.00) | 0.996 | 0.740 | 0.995 | 0.769 | 0.849 | 0.764 | 0.931 |
| IND | IBD | CDW | Microscopic exam | 0.024 | 0.730 (0.63 - 0.82) | 1.000 (0.96 - 1.00) | 0.991 | 1.000 | 0.993 | 0.844 | 0.995 | 0.852 | 0.781 |
| IND | IBD | MVP | Full pathology report | 0.019 | 0.903 (0.83 - 0.96) | 0.991 (0.96 - 1.00) | 0.999 | 0.674 | 0.998 | 0.772 | 0.805 | 0.775 | 0.935 |

**Supplementary Table S3: Llama-3-8B-Instruct results.** Llama-3 showed similar results to Gemma-2, but underperformed slightly in most cases. 95% confidence intervals for PPV and NPV were approximated using the binomial distribution (see Supplementary Methods section *Calculating performance metrics*). Shading progression has lower value (red) = 0.5, middle value (white) = 0.9 and upper value (green) = 1. CRC = Invasive colorectal cancer (invasive colorectal adenocarcinoma); HGD/CRC = High-grade dysplasia and/or adenocarcinoma; IND = Indefinite for dysplasia; IBD = Inflammatory bowel disease; PPV = Positive predictive value; NPV = Negative predictive value; LB = Lower bound; UB = Upper bound; PPVc = Calibrated positive predictive value (calibrated precision)[15]. F1c = Calibrated F1 score[15]. MCC = Matthew's correlation coefficient.

| Task | Model | Model prevalence estimate | PPV (LB - UB) | NPV (LB - UB) | PPVc | Recall (Sensitivity) | Specificity | F1 | F1c | MCC | Cohen's kappa |
|---|---|---|---|---|---|---|---|---|---|---|---|
| CRC | llama3.2_3B | 0.221 | 0.941 (0.88 - 0.98) | 0.925 (0.88 - 0.96) | 0.986 | 0.779 | 0.982 | 0.852 | 0.871 | 0.812 | 0.889 |
| CRC | gemma2_2B | 0.237 | 0.938 (0.88 - 0.98) | 0.942 (0.89 - 0.97) | 0.983 | 0.835 | 0.980 | 0.883 | 0.903 | 0.847 | 0.889 |
| CRC | either_small | 0.260 | 0.925 (0.87 - 0.97) | 0.965 (0.92 - 0.99) | 0.975 | 0.902 | 0.973 | 0.913 | 0.937 | 0.883 | 0.889 |
| CRC | both_small | 0.198 | 0.958 (0.91 - 0.99) | 0.904 (0.85 - 0.94) | 0.992 | 0.712 | 0.989 | 0.817 | 0.829 | 0.777 | 0.889 |
| HGD/CRC | llama3.2_3B | 0.156 | 0.866 (0.76 - 0.97) | 0.955 (0.91 - 0.98) | 0.978 | 0.782 | 0.975 | 0.822 | 0.869 | 0.788 | 0.947 |
| HGD/CRC | gemma2_2B | 0.188 | 0.828 (0.72 - 0.94) | 0.979 (0.94 - 1.00) | 0.959 | 0.900 | 0.961 | 0.862 | 0.928 | 0.833 | 0.947 |
| HGD/CRC | either_small | 0.199 | 0.804 (0.70 - 0.92) | 0.985 (0.95 - 1.00) | 0.947 | 0.929 | 0.953 | 0.862 | 0.938 | 0.834 | 0.947 |
| HGD/CRC | both_small | 0.145 | 0.901 (0.80 - 0.99) | 0.950 (0.91 - 0.98) | 0.986 | 0.753 | 0.983 | 0.821 | 0.854 | 0.792 | 0.947 |
| Dysplasia | llama3.2_3B | 0.154 | 0.947 (0.87 - 1.00) | 0.913 (0.86 - 0.95) | 0.993 | 0.665 | 0.990 | 0.782 | 0.797 | 0.751 | 0.920 |
| Dysplasia | gemma2_2B | 0.212 | 0.906 (0.82 - 0.98) | 0.966 (0.93 - 0.99) | 0.976 | 0.878 | 0.974 | 0.892 | 0.925 | 0.862 | 0.920 |
| Dysplasia | either_small | 0.222 | 0.886 (0.80 - 0.97) | 0.971 (0.93 - 0.99) | 0.968 | 0.898 | 0.968 | 0.892 | 0.932 | 0.861 | 0.920 |
| Dysplasia | both_small | 0.144 | 0.980 (0.93 - 1.00) | 0.909 (0.86 - 0.95) | 0.998 | 0.646 | 0.996 | 0.778 | 0.784 | 0.756 | 0.920 |
| IND | llama3.2_3B | 0.019 | 0.902 (0.84 - 0.95) | 0.992 (0.97 - 1.00) | 0.999 | 0.684 | 0.998 | 0.778 | 0.812 | 0.781 | 0.931 |
| IND | gemma2_2B | 0.019 | 0.902 (0.83 - 0.95) | 0.992 (0.97 - 1.00) | 0.999 | 0.678 | 0.998 | 0.774 | 0.808 | 0.777 | 0.931 |
| IND | either_small | 0.020 | 0.892 (0.83 - 0.94) | 0.993 (0.97 - 1.00) | 0.998 | 0.715 | 0.998 | 0.794 | 0.833 | 0.794 | 0.931 |
| IND | both_small | 0.018 | 0.913 (0.85 - 0.96) | 0.991 (0.96 - 1.00) | 0.999 | 0.647 | 0.998 | 0.758 | 0.786 | 0.764 | 0.931 |

**Supplementary Table S4: Small models (Llama-3.2-3B and Gemma-2-2B) struggle with recall.** Small models were applied to the IBD-colitis validation set in MVP. Gemma-2-2B outperforms Llama-3.2-3B. 95% confidence intervals for PPV and NPV were approximated using the binomial distribution (see Supplementary Methods section *Calculating performance metrics*). CRC = Invasive colorectal cancer (invasive colorectal adenocarcinoma); HGD/CRC = High-grade dysplasia or adenocarcinoma; IND = Indefinite for dysplasia; IBD = Inflammatory bowel disease; PPV = Positive predictive value; NPV = Negative predictive value; LB = Lower bound; UB = Upper bound; $PPV_c$ = Calibrated positive predictive value (calibrated precision). $F1_c$ = Calibrated F1 score[15]. MCC = Matthew's correlation coefficient.

| Task | Cohort | Source | Model | Input | Model prevalence estimate | PPV (LB - UB) | NPV (LB - UB) | PPVc | Recall (Sensitivity) | Specificity | F1 | F1c | MCC | Cohen's kappa |
|---|---|---|---|---|---|---|---|---|---|---|---|---|---|---|
| CRC | IBD | MVP | both | Microscopic exam | 0.245 | 0.966 (0.92 - 0.99) | 0.962 (0.92 - 0.99) | 0.990 | 0.891 | 0.989 | 0.927 | 0.938 | 0.904 | 0.889 |
| CRC | IBD | MVP | either | Microscopic exam | 0.284 | 0.937 (0.89 - 0.98) | 1.000 (0.98 - 1.00) | 0.974 | 1.000 | 0.975 | 0.967 | 0.987 | 0.956 | 0.889 |
| CRC | IBD | MVP | both | Full pathology report | 0.251 | 0.895 (0.81 - 0.96) | 0.941 (0.88 - 0.98) | 0.968 | 0.835 | 0.964 | 0.864 | 0.897 | 0.817 | 0.877 |
| CRC | IBD | MVP | either | Full pathology report | 0.311 | 0.822 (0.73 - 0.91) | 0.981 (0.93 - 1.00) | 0.915 | 0.950 | 0.924 | 0.881 | 0.932 | 0.838 | 0.877 |
| CRC | IBD | CDW | both | Microscopic exam | 0.249 | 0.979 (0.93 - 1.00) | 0.970 (0.92 - 0.99) | 0.994 | 0.915 | 0.993 | 0.946 | 0.953 | 0.928 | 0.910 |
| CRC | IBD | CDW | either | Microscopic exam | 0.277 | 0.909 (0.83 - 0.98) | 0.980 (0.93 - 1.00) | 0.965 | 0.945 | 0.966 | 0.926 | 0.955 | 0.899 | 0.910 |
| CRC | non-IBD | MVP | both | Microscopic exam | 0.505 | 0.986 (0.95 - 1.00) | 0.875 (0.81 - 0.93) | 0.988 | 0.890 | 0.984 | 0.935 | 0.936 | 0.868 | 0.920 |
| CRC | non-IBD | MVP | either | Microscopic exam | 0.538 | 0.975 (0.94 - 0.99) | 0.923 (0.87 - 0.96) | 0.973 | 0.936 | 0.969 | 0.955 | 0.954 | 0.902 | 0.920 |
| CRC | non-IBD | CDW | both | Microscopic exam | 0.519 | 0.979 (0.93 - 1.00) | 0.920 (0.85 - 0.97) | 0.979 | 0.929 | 0.976 | 0.954 | 0.954 | 0.902 | 0.940 |
| CRC | non-IBD | CDW | either | Microscopic exam | 0.558 | 0.963 (0.91 - 0.99) | 0.979 (0.93 - 1.00) | 0.954 | 0.983 | 0.954 | 0.973 | 0.968 | 0.940 | 0.940 |
| HGD/CRC | IBD | MVP | both | Microscopic exam | 0.146 | 0.973 (0.93 - 0.99) | 0.964 (0.93 - 0.99) | 0.996 | 0.824 | 0.995 | 0.892 | 0.902 | 0.876 | 0.947 |
| HGD/CRC | IBD | MVP | either | Microscopic exam | 0.183 | 0.911 (0.82 - 0.98) | 0.993 (0.96 - 1.00) | 0.979 | 0.967 | 0.980 | 0.938 | 0.973 | 0.926 | 0.947 |
| HGD/CRC | IBD | MVP | both | Full pathology report | 0.141 | 0.923 (0.82 - 0.99) | 0.961 (0.92 - 0.99) | 0.989 | 0.795 | 0.987 | 0.854 | 0.882 | 0.832 | 0.941 |
| HGD/CRC | IBD | MVP | either | Full pathology report | 0.198 | 0.792 (0.67 - 0.92) | 0.991 (0.95 - 1.00) | 0.942 | 0.958 | 0.951 | 0.867 | 0.950 | 0.844 | 0.941 |
| HGD/CRC | IBD | CDW | both | Microscopic exam | 0.180 | 0.979 (0.93 - 1.00) | 0.960 (0.90 - 0.99) | 0.996 | 0.844 | 0.995 | 0.907 | 0.914 | 0.888 | 0.970 |
| HGD/CRC | IBD | CDW | either | Microscopic exam | 0.219 | 0.920 (0.84 - 0.98) | 0.990 (0.94 - 1.00) | 0.977 | 0.961 | 0.978 | 0.940 | 0.969 | 0.924 | 0.970 |
| HGD/CRC | non-IBD | MVP | both | Microscopic exam | 0.260 | 0.993 (0.96 - 1.00) | 0.927 (0.87 - 0.97) | 0.998 | 0.826 | 0.997 | 0.902 | 0.904 | 0.871 | 0.947 |
| HGD/CRC | non-IBD | MVP | either | Microscopic exam | 0.304 | 0.978 (0.94 - 1.00) | 0.979 (0.94 - 1.00) | 0.991 | 0.953 | 0.990 | 0.965 | 0.971 | 0.950 | 0.947 |
| HGD/CRC | non-IBD | CDW | both | Microscopic exam | 0.370 | 1.000 (0.96 - 1.00) | 0.970 (0.91 - 1.00) | 1.000 | 0.951 | 1.000 | 0.975 | 0.975 | 0.961 | 0.950 |
| HGD/CRC | non-IBD | CDW | either | Microscopic exam | 0.389 | 0.984 (0.93 - 1.00) | 0.990 (0.94 - 1.00) | 0.990 | 0.984 | 0.990 | 0.984 | 0.987 | 0.973 | 0.950 |
| Dysplasia | IBD | MVP | both | Microscopic exam | 0.212 | 0.980 (0.94 - 1.00) | 0.987 (0.95 - 1.00) | 0.995 | 0.952 | 0.995 | 0.966 | 0.973 | 0.957 | 0.920 |
| Dysplasia | IBD | MVP | either | Microscopic exam | 0.224 | 0.951 (0.90 - 0.99) | 0.993 (0.96 - 1.00) | 0.986 | 0.976 | 0.986 | 0.963 | 0.981 | 0.953 | 0.920 |
| Dysplasia | IBD | MVP | both | Full pathology report | 0.207 | 1.000 (0.97 - 1.00) | 0.987 (0.94 - 1.00) | 1.000 | 0.953 | 1.000 | 0.976 | 0.976 | 0.970 | 0.967 |
| Dysplasia | IBD | MVP | either | Full pathology report | 0.220 | 0.954 (0.89 - 1.00) | 0.992 (0.95 - 1.00) | 0.987 | 0.970 | 0.987 | 0.962 | 0.978 | 0.951 | 0.967 |
| Dysplasia | IBD | CDW | both | Microscopic exam | 0.304 | 0.980 (0.93 - 1.00) | 0.990 (0.95 - 1.00) | 0.991 | 0.977 | 0.991 | 0.978 | 0.984 | 0.969 | 0.950 |
| Dysplasia | IBD | CDW | either | Microscopic exam | 0.324 | 0.940 (0.88 - 0.99) | 1.000 (0.96 - 1.00) | 0.971 | 1.000 | 0.972 | 0.969 | 0.985 | 0.956 | 0.950 |
| Dysplasia | non-IBD | MVP | both | Microscopic exam | 0.895 | 0.987 (0.95 - 1.00) | 0.973 (0.93 - 0.99) | 0.897 | 0.997 | 0.895 | 0.992 | 0.944 | 0.926 | 0.953 |
| Dysplasia | non-IBD | MVP | either | Microscopic exam | 0.898 | 0.986 (0.95 - 1.00) | 0.993 (0.96 - 1.00) | 0.889 | 0.999 | 0.889 | 0.993 | 0.941 | 0.933 | 0.953 |
| Dysplasia | non-IBD | CDW | both | Microscopic exam | 0.875 | 0.990 (0.95 - 1.00) | 0.980 (0.93 - 1.00) | 0.934 | 0.997 | 0.933 | 0.994 | 0.965 | 0.950 | 0.920 |
| Dysplasia | non-IBD | CDW | either | Microscopic exam | 0.879 | 0.987 (0.95 - 1.00) | 0.990 (0.94 - 1.00) | 0.914 | 0.999 | 0.914 | 0.993 | 0.955 | 0.944 | 0.920 |
| IND | IBD | MVP | both | Microscopic exam | 0.021 | 0.868 (0.80 - 0.92) | 0.993 (0.97 - 1.00) | 0.998 | 0.728 | 0.997 | 0.791 | 0.841 | 0.790 | 0.931 |
| IND | IBD | MVP | either | Microscopic exam | 0.036 | 0.692 (0.44 - 0.90) | 1.000 (0.98 - 1.00) | 0.984 | 1.000 | 0.989 | 0.818 | 0.992 | 0.827 | 0.931 |
| IND | IBD | MVP | both | Full pathology report | 0.019 | 0.921 (0.85 - 0.97) | 0.991 (0.96 - 1.00) | 0.999 | 0.674 | 0.998 | 0.779 | 0.805 | 0.783 | 0.935 |
| IND | IBD | MVP | either | Full pathology report | 0.029 | 0.628 (0.39 - 0.90) | 0.992 (0.96 - 1.00) | 0.988 | 0.696 | 0.989 | 0.660 | 0.817 | 0.652 | 0.935 |
| IND | IBD | CDW | both | Microscopic exam | 0.022 | 0.753 (0.65 - 0.84) | 0.999 (0.97 - 1.00) | 0.993 | 0.959 | 0.994 | 0.843 | 0.976 | 0.847 | 0.781 |
| IND | IBD | CDW | either | Microscopic exam | 0.034 | 0.520 (0.30 - 0.79) | 1.000 (0.96 - 1.00) | 0.969 | 1.000 | 0.983 | 0.684 | 0.984 | 0.715 | 0.781 |

**Supplementary Table S5: Requiring either or both Llama-3-8B and Gemma-2-9B to answer "Yes" achieves similar results to using just one model.** 95% confidence intervals for PPV and NPV were approximated using the binomial distribution (see Supplementary Methods section *Calculating performance metrics*). CRC = Invasive colorectal cancer (invasive colorectal adenocarcinoma); HGD/CRC = High-grade dysplasia or adenocarcinoma; IND = Indefinite for dysplasia; IBD = Inflammatory bowel disease; PPV = Positive predictive value; NPV = Negative predictive value; LB = Lower bound; UB = Upper bound; PPVc = Calibrated positive predictive value (calibrated precision). F1c = Calibrated F1 score[15]. MCC = Matthew's correlation coefficient.

| Task (all MVP) | LLMs run with full note and Microscopic Exam separately | Either LLM disagrees with itself seeing full note compared to seeing Microscopic Exam | Validation using full note disagrees with initial validation using Microscopic Exam |
|---|---|---|---|
| invasive crc | 227 | 15 | 1 |
| hgd and/or crc | 237 | 26 | 1 |
| any dysplasia | 239 | 4 | 0 |
| indefinite for dysplasia | 253 | 23 | 3 |

**Supplementary Table S6: Diagnoses differing between full note and microscopic exam section.** When doing validation for the full pathology reports, we re-validated only the reports where the LLM response changed. For example, if both Llama-3 and Gemma-2 respond "Yes" when given the Microscopic Exam section but one of them responds "No" when given the full pathology report, we would re-validate that report using the full pathology report. Otherwise, if the answers stayed the same, we would use the Microscopic Exam validation as the full Pathology report validation. It is possible that the full pathology report contains information missing from the Microscopic Exam section due to the way the note is written and the way the Pathology Domain is created. For our full note evaluation, we selected one full pathology report from the linked match for each pathology report in the Pathology Domain. If multiple notes matched a single pathology report, the note was chosen randomly. If there were no notes matching a given pathology report, the report was removed from consideration for the full pathology report evaluation.

| Study | Approach | Task | Cohort | Input type | Number validated | Number of model positives validated | Number of model negatives validated | F1 | Notes |
|---|---|---|---|---|---|---|---|---|---|
| This work (gemma-2) | LLM + rules-based | any dysplasia | No IBD-colitis in VA (CDW) | Microscopic exam | 200 | 103 | 97 | 0.993 | |
| This work (gemma-2) | LLM + rules-based | any dysplasia | No IBD-colitis in VA (MVP) | Microscopic exam | 300 | 154 | 146 | 0.992 | |
| This work (llama-3) | LLM + rules-based | any dysplasia | No IBD-colitis in VA (CDW) | Microscopic exam | 200 | 100 | 100 | 0.994 | |
| This work (llama-3) | LLM + rules-based | any dysplasia | No IBD-colitis in VA (MVP) | Microscopic exam | 300 | 150 | 150 | 0.992 | |
| Bae (2022) | Rules-based | Conventional adenoma | general population in South Korea | Full pathology report | 1000 | | | 0.99 | |
| Benson (2023) | Rules-based | "across all features" | general population in U.S. | Full pathology report | 150 | | | 0.984 | F1-score not reported for identifying adenoma. |
| Carrell (2017) | Rules-based | Any adenoma | general population in U.S. (40+, outpatient) | Full pathology report | 1469 | | | 0.95 | |
| Fevrier (2020) | Rules-based | Adenoma | general population in U.S. (50+) | Full pathology report | 100 | | | 0.98 | F1-score not reported. Calculated based on PPV and recall reported. |
| Gupta (2023) | Rules-based | Any adenoma | non-IBD in VA | Full pathology report | 200 | | | 1 | F1-score not reported. Perfect NPV and PPV. |
| Harkema (2011) | Rules-based | "adenomatous" | general population in U.S. | Full pathology report | 226 | | | 0.98 | |
| Imler (2013) | Rules-based | Tubular adenoma | non-IBD in VA (40+) | Full pathology report | 350 | | | 0.98 | 500 total annotated. 150 used for training, 350 as true test set. F1-score reported for "most advanced lesion". Calculated based on precision and recall reported for tubular adenoma. |
| Nayor (2018) | Rules-based | Adenoma | general population in U.S. | Full pathology report | 100 | 69 | 31 | 1 | |
| Raju (2015) | Rules-based | Adenoma | non-IBD in U.S. (50-75, first screening colonoscopy) | Full pathology report | 2259 | 962 | 1297 | 0.996 | F1-score calculated based on confusion matrix given in table 3. |
| Syed (2022) | Deep-learning | "neoplastic polyps" | general population in U.S. | Full pathology report | 219 | 45 | 174 | 0.95 | |
| This work (gemma-2) | LLM + rules-based | hgd and/or crc | No IBD-colitis in VA (CDW) | Microscopic exam | 200 | 103 | 97 | 0.975 | |
| This work (gemma-2) | LLM + rules-based | hgd and/or crc | No IBD-colitis in VA (MVP) | Microscopic exam | 300 | 159 | 141 | 0.965 | |
| This work (llama-3) | LLM + rules-based | hgd and/or crc | No IBD-colitis in VA (CDW) | Microscopic exam | 200 | 100 | 100 | 0.975 | |
| This work (llama-3) | LLM + rules-based | hgd and/or crc | No IBD-colitis in VA (MVP) | Microscopic exam | 300 | 150 | 150 | 0.902 | |
| Bae (2022) | Rules-based | "advanced adenoma" | general population in South Korea | Full pathology report | 1000 | | | 0.99 | "Advanced adenomas were defined as adenomas ≥1 cm in size or with pathological features such as high-grade dysplasia or villous features." |
| Gupta (2023) | Rules-based | Adenoma with HGD | non-IBD in VA | Full pathology report | 200 | 100 | 100 | 0.99 | F1-score calculated from given PPV and NPV assuming an "adenoma with HGD" prevalence of 3.9% from same study |
| Gupta (2023) | Rules-based | Carcinoma in situ | non-IBD in VA | Full pathology report | 200 | 100 | 100 | 0.757 | F1-score calculated from given PPV and NPV assuming an estimated prevalence of 3.9% (prevalence of adenoma with HGD). Likely overestimate of prevalence and F1-score. |
| Harkema (2011) | Rules-based | "bad pathology" | general population in U.S. | Full pathology report | 226 | 16 | 210 | 0.94 | "Whether any adenoma has villous component, high-grade dysplasia, or pathology shows invasive cancer" |
| Imler (2013) | Rules-based | "advanced adenoma" | non-IBD in VA (40+) | Full pathology report | 350 | | | 0.915 | F1-score calculated based on precision and recall given. "Advanced adenomas were defined as those with villous features, carcinoma in situ, high-grade dysplasia, or adenoma with size on colonoscopy report ≤10 mm with size determined by the endoscopist." |
| This work (gemma-2) | LLM + rules-based | invasive crc | No IBD-colitis in VA (CDW) | Microscopic exam | 200 | 102 | 98 | 0.964 | |
| This work (gemma-2) | LLM + rules-based | invasive crc | No IBD-colitis in VA (MVP) | Microscopic exam | 300 | 154 | 146 | 0.950 | |
| This work (llama-3) | LLM + rules-based | invasive crc | No IBD-colitis in VA (CDW) | Microscopic exam | 200 | 100 | 100 | 0.959 | |
| This work (llama-3) | LLM + rules-based | invasive crc | No IBD-colitis in VA (MVP) | Microscopic exam | 300 | 150 | 150 | 0.939 | |
| Gupta (2023) | Rules-based | Adenocarcinoma | non-IBD in VA | Full pathology report | 200 | 100 | 100 | 0.867 | F1-score calculated from given PPV and NPV assuming an estimated prevalence of 5%. Likely overestimate of prevalence and F1-score. |
| Imler (2013) | Rules-based | Carcinoma | non-IBD in VA (40+) | Full pathology report | 350 | 10 (estimated) | 340 | 0.947 | Assuming 10 positive cases validated based on low prevalence and exactly 90.0% reported PPV. F1-score calculated based on precision and recall reported |
| Nayor (2018) | Rules-based | Adenocarcinoma | general population in U.S. | Full pathology report | 100 | 2 | 98 | 1 | |
| Syed (2022) | Deep-learning | Malignant carcinoma | general population in U.S. | Full pathology report | 219 | 17 | 202 | 0.937 | |

**Supplementary Table S7: Comparison to previously published approaches[16–25].** Comparing our validation set (non-IBD) with previously published studies in predominantly non-IBD patient cohorts. All perform well in identifying common features such as any adenoma or dysplasia. Rarer features such as HGD and CRC show reduced performance and/or small validated sample sizes. When F1-score was not reported, it was calculated, if possible, from the recall and PPV, adjusting for prevalence where necessary (see notes column). Analyses from different publications differed slightly in their methods, but we compared similar approaches where possible. Sample sizes below 30 highlighted in red.

9. Gemma Team, Thomas Mesnard, Cassidy Hardin, et al. Gemma. doi:10.34740/KAGGLE/M/3301

**Million Veteran Program:**
**Core Acknowledgements for Publications**
**May 2024**

### MVP Program Office

- Sumitra Muralidhar, Ph.D., Program Director
  US Department of Veterans Affairs, 810 Vermont Avenue NW, Washington, DC 20420
- Jennifer Moser, Ph.D., Associate Director, Scientific Programs
  US Department of Veterans Affairs, 810 Vermont Avenue NW, Washington, DC 20420
- Jennifer E. Deen, B.S., Associate Director, Cohort & Public Relations
  US Department of Veterans Affairs, 810 Vermont Avenue NW, Washington, DC 20420

### MVP Executive Committee

- Co-Chair: Philip S. Tsao, Ph.D.
  VA Palo Alto Health Care System, 3801 Miranda Avenue, Palo Alto, CA 94304
- Co-Chair: Sumitra Muralidhar, Ph.D.
  US Department of Veterans Affairs, 810 Vermont Avenue NW, Washington, DC 20420
- J. Michael Gaziano, M.D., M.P.H.
  VA Boston Healthcare System, 150 S. Huntington Avenue, Boston, MA 02130
- Elizabeth Hauser, Ph.D.
  Durham VA Medical Center, 508 Fulton Street, Durham, NC 27705
- Amy Kilbourne, Ph.D., M.P.H.
  VA HSR&D, 2215 Fuller Road, Ann Arbor, MI 48105
- Michael Matheny, M.D., M.S., M.P.H.
  VA Tennessee Valley Healthcare System, 1310 24th Ave. South, Nashville, TN 37212
- Dave Oslin, M.D.
  Philadelphia VA Medical Center, 3900 Woodland Avenue, Philadelphia, PA 19104
- Deepak Voora, MD
  Durham VA Medical Center, 508 Fulton Street, Durham, NC 27705

### MVP Co-Principal Investigators

- J. Michael Gaziano, M.D., M.P.H.
  VA Boston Healthcare System, 150 S. Huntington Avenue, Boston, MA 02130
- Philip S. Tsao, Ph.D.
  VA Palo Alto Health Care System, 3801 Miranda Avenue, Palo Alto, CA 94304

### MVP Core Operations

- Jessica V. Brewer, M.P.H., Director, MVP Cohort Operations
  VA Boston Healthcare System, 150 S. Huntington Avenue, Boston, MA 02130
- Mary T. Brophy M.D., M.P.H., Director, VA Central Biorepository
  VA Boston Healthcare System, 150 S. Huntington Avenue, Boston, MA 02130
- Kelly Cho, M.P.H, Ph.D., Director, MVP Phenomics

VA Boston Healthcare System, 150 S. Huntington Avenue, Boston, MA 02130

- Lori Churby, B.S., Director, MVP Regulatory Affairs
  VA Palo Alto Health Care System, 3801 Miranda Avenue, Palo Alto, CA 94304
- Scott L. DuVall, Ph.D., Director, VA Informatics and Computing Infrastructure (VINCI)
  VA Salt Lake City Health Care System, 500 Foothill Drive, Salt Lake City, UT 84148
- Saiju Pyarajan Ph.D., Director, Data and Computational Sciences
  VA Boston Healthcare System, 150 S. Huntington Avenue, Boston, MA 02130
- Robert Ringer, Pharm.D., Director, VA Albuquerque Central Biorepository
  New Mexico VA Health Care System, 1501 San Pedro Drive SE, Albuquerque, NM 87108
- Luis E. Selva, Ph.D., Director, MVP Biorepository Coordination
  VA Boston Healthcare System, 150 S. Huntington Avenue, Boston, MA 02130
- Shahpoor (Alex) Shayan, M.S., Director, MVP PRE Informatics
  VA Boston Healthcare System, 150 S. Huntington Avenue, Boston, MA 02130
- Brady Stephens, M.S., Principal Investigator, MVP Information Center
  Canandaigua VA Medical Center, 400 Fort Hill Avenue, Canandaigua, NY 14424
- Stacey B. Whitbourne, Ph.D., Director, MVP Cohort Development and Management
  VA Boston Healthcare System, 150 S. Huntington Avenue, Boston, MA 02130

**MVP Publications and Presentations Committee**

- Co-Chair: Themistocles L. Assimes, M.D., Ph. D
  VA Palo Alto Health Care System, 3801 Miranda Avenue, Palo Alto, CA 94304
- Co-Chair: Adriana Hung, M.D.; M.P.H
  VA Tennessee Valley Healthcare System, 1310 24th Ave. South, Nashville, TN 37212
- Co-Chair: Henry Kranzler, M.D.

Philadelphia VA Medical Center, 3900 Woodland
